# Advancing maternal and newborn health care measurement: Developing quality of care indices for postnatal and small and/or sick newborn care in low- and middle-income countries

**DOI:** 10.1101/2024.10.03.24314852

**Authors:** Ashley Sheffel, Shannon King, Louise Tina Day, Tanya Marchant, Moise Muzigaba, Jennifer Requejo, Emily Carter, Melinda K. Munos

**Author notes:** **Corresponding author:** Dr. Ashley Sheffel, DrPH Johns Hopkins University Bloomberg School of Public Health, Department of International Health 615 N Wolfe St. Baltimore, Maryland, USA 21205-2103.

## Abstract

**Background:** High-quality healthcare for pregnant women and newborns, particularly postnatal care (PNC) and small and/or sick newborn care (SSNC), is essential to reducing maternal and newborn morbidity and mortality in low- and middle-income countries (LMICs). Poor quality of care is a major contributor to preventable morbidity and mortality, emphasizing the need for improvements in health service delivery, which requires measuring and monitoring quality of care (QoC). Although indicators measuring QoC have been identified, there is a current gap in the availability of composite indicators that can summarize the complex, multidimensional nature of QoC. This study systematically developed three composite QoC indices for maternal PNC, newborn PNC, and SSNC feasible to measure using existing data in LMICs.

**Methods:** A four-step process was used to define the indices: (1) Intervention selection: Key interventions were identified by reviewing global clinical guidelines and QoC frameworks; (2) Guideline review and item identification: Discrete items recommended for delivery of each of the selected interventions were extracted from intervention-specific guidelines; (3) Data mapping: These items were mapped to health facility survey data to assess their alignment with standardized tools; and (4) Final index development: A quality readiness index (QRI) was developed for each service area based on QoC frameworks, available data, and clinical guidelines.

**Results:** The maternal PNC-QRI includes 12 interventions and contains 24 items. The newborn PNC-QRI includes 3 interventions and contains 16 items. The SSNC-QRI includes 8 interventions and contains 48 items. Data gaps for maternal PNC, newborn PNC, and SSNC led to the exclusion of some evidence-based interventions and limited item inclusion. No data on provision/experience of care were available for PNC or SSNC, thus the indices reflect only facility readiness.

**Conclusions:** The three QRIs developed provide composite measures for PNC and SSNC readiness and can be adapted at country level and operationalized using health facility assessment survey data, facilitating their use by decision-makers for planning and resource allocation. Revision of existing health facility assessments to address gaps in readiness and provision/experience of care measurement for PNC and SSNC would bolster efforts to monitor and improve QoC for mothers and newborns.

## BACKGROUND

High quality health services are crucial to achieving maternal, newborn and child health (MNCH) goals, including Sustainable Development Goal 3 (SDG) which aims to ensure healthy lives and promote well-being for everyone at all ages [1] [2]. Recognizing the importance of these services, there has been an increasing emphasis on improving, measuring, and monitoring both access to and the quality of MNCH health services. A significant initiative in this regard is the Every Newborn Action Plan (ENAP), launched in 2014, which is a comprehensive, multi-partner effort calling on stakeholders to improve access and quality of care (QoC) for all pregnant women and newborns. This initiative underscores the need for enhanced measurement, particularly concerning quality of care, which is a strategic objective of ENAP [3]. However, there is still a lack of standardized indicators for effectively monitoring the quality of maternal and newborn care thus service contact coverage indicators remain common for monitoring progress in service delivery.

While service contact coverage indicators, such as postnatal care contacts for mothers and newborns within two days of delivery, provide valuable information on access to health services, previous research has shown the contact-content coverage gap — these indicators do not capture the specific interventions delivered or quality of care provided during the service contact [4,5]. Monitoring efforts for maternal and newborn health have revealed a concerning trend: despite substantial improvements in service contact coverage, many countries are not achieving rapid reductions in maternal and newborn mortality [6]. This finding underscores that providing and monitoring high-quality maternal and newborn health services including small and/or sick newborn care (SSNC) (small newborns being those that weigh <2500g at birth; sick newborns being those that have any medical or surgical conditions during the neonatal period (days 0-28) among babies of all birthweights [7]), and routine postnatal care (PNC) (care for mothers and newborns beginning immediately after birth and extending up to six weeks after birth [8]) is critical to reducing maternal and neonatal morbidity and mortality in low- and middle-income countries (LMICs).

Measuring maternal and newborn quality of care requires a clear definition of QoC, along with standardized indicators and data sources for operationalization. The World Health Organization (WHO) has developed a definition for “quality of care” — ‘The extent to which health care services provided to individuals and patient populations improve desired health outcomes. In order to achieve this, health care needs to be safe, effective, timely, efficient, equitable, and people-centred.’ — and a framework for improving the QoC for mothers and newborns around the time of childbirth [9]. In addition, the WHO has published standards for improving quality of maternal and newborn care in health facilities, including for small and/or sick newborns, which contain quality standards, quality statements, and quality measures (350 quality measures for maternal and newborn health; 578 quality measures for small and/or sick newborn care) [7,10]. The maternal and newborn health QoC monitoring framework recognizes stakeholders have different QoC measurement needs and proposes several measurement components including a core set of indicators (a small set of prioritized input, process, outcome, and impact indicators to track and compare across and within regions and countries) and a quality improvement indicator catalogue (a menu of indicators to support quality improvement at facility and subnational levels) [11].

Although indicators measuring quality of care have been identified, there is a current gap in the availability of composite indicators that can summarize the complex, multidimensional nature of QoC. Composite indicators are formed when individual indicators are combined into a single index which can be useful for assessing and monitoring overall health system progress and benchmarking within and across countries [12–14]. Composite indicators of service quality are particularly useful for measuring and tracking effective coverage (EC) - the proportion of a population in need of a service that received the service with sufficient quality to achieve a positive health outcome-through the use of effective coverage cascades. One common approach to estimating EC is to link composite indicators for service readiness and process quality to measures of service contact coverage [4,15–18]. Utilizing existing data generated from commonly implemented health facility assessments (HFAs) in LMICs, such as the Service Provision Assessment (SPA), Service Availability and Readiness Assessment (SARA), and Harmonized Health Facility Assessment (HHFA), which are designed to assess the quality of services, provides an efficient, sustainable way to measure both composite quality indicators and support effective coverage measurement [19–22]. Given the need for composite indicators that can be measured with data currently available in LMICs, this study aimed to systematically develop QoC indices for maternal PNC, newborn PNC, and SSNC using existing HFA data.

## METHODS

To define QoC indices for PNC for women, PNC for newborns, and SSNC in LMICs in LMICs we used a four-step process, similar to the approach previously taken for developing QoC indices for maternal nutrition [23]:

1. **Intervention selection:** We reviewed global clinical guidelines and QoC frameworks to select recommended interventions.
2. **Guideline review and item identification:** We reviewed intervention-specific clinical and service implementation guidelines to identify discrete elements or ‘items’ recommended for delivery of each of the selected interventions using the WHO MNH QoC framework as an organizing framework.
3. **Data mapping:** We matched the identified discrete items to available health facility survey data, assessing the degree of alignment with standardized health facility assessments.
4. **Final index development:** We developed final QoC indices for each service area informed by QoC frameworks, clinical guidelines, and data availability.

The SSNC index development process was funded through a separate mechanism from the PNC work, with more limited objectives, focusing only on readiness and not provision/experience of care.

Therefore, for SSNC, the above process was implemented only for readiness whereas for maternal and newborn PNC, the process was implemented for readiness and provision/experience of care. We also note that large HFA programs including the SPA, SARA, and HHFA do not currently collect provision or experience of care data for SSNC.

### Step 1: Intervention selection

Maternal and newborn PNC interventions were identified through a review of the 2022 WHO recommendations on maternal and newborn care for a positive postnatal experience (hereafter “WHO PNC guidelines” [8]. Interventions for SSNC were identified through a review of WHO guidelines and previous studies assessing facility readiness for SSNC [7,24–26].

### Step 2: Guideline review and item identification

PNC interventions were included in guideline review and item identification if WHO recommended the intervention either for all or for specific contexts and if it was a clinical intervention. PNC interventions were excluded if the intervention was not recommended by WHO or was a best practice rather than a clinical intervention. SSNC interventions were included in guideline review and item identification if they were routine and essential newborn care or special newborn care clinical interventions. SSNC interventions were excluded if they were recommended at intensive care level or transition to intensive newborn care as these are highly specialized services, which are not collected in the SPA/SARA/HHFA, and our aim was to use existing data.

The aim of the guideline extraction step was to identify discrete elements or ‘items’ recommended for delivery of each of the selected interventions. For each intervention that met the inclusion criteria, we first reviewed WHO facility-level service delivery guidelines; where those were lacking, we hand-searched the references from the key documents used to identify interventions in step 1 and identified and reviewed other available guidance and protocols (e.g., Médecins Sans Frontières, the American Academy of Pediatrics, country-specific guidelines), and/or published peer reviewed literature (see **Table S2** in the Online Supplementary Document for more details). Guideline extraction was organized by the quality domains proposed by the WHO Quality of Care Framework for Maternal and Newborn Health – provision of care, experience of care, and service readiness (see ***Box 1*** for definitions) [10,27]. Provision of care was further categorized into the sub-domains of assessment, intervention, and documentation and referral. Service readiness was further categorized into the sub-domains of basic amenities, equipment and supplies, medicines and commodities, diagnostics, guidelines and staff training, and, for SSNC only, routine service. The guideline extraction process was conducted by two researchers at Johns Hopkins University (SK and AS).

### Step 3: Data mapping

The SPA, SARA, and HHFA are three of the most widely implemented HFAs in LMICs and provide nationally representative data on health service delivery including service readiness across the continuum of care and provision/experience of care for select services [28–30]. Developing QoC indices using the data available from these surveys provides a means to operationalize the QoC indices using existing data in LMICs. Each item identified during the guideline extraction process was matched with available items from the SPA and SARA standard questionnaires. Both SPA and SARA were updated in 2022, with the SARA replaced by the HHFA [19–22,31]. As such, we mapped to the older questionnaires, which correspond to existing country data, as well as the newer questionnaires, which represent what data will be available from future country surveys. The level of agreement between the item in the guideline and item in the HFA questionnaire was classified as an exact match, high/low partial match, or nonmatch (**Box 2**). All items that were an exact match, high partial match, or low partial match were eligible for inclusion in the QoC indices.

### Step 4: Final index development

PNC and SSNC interventions were excluded in final index development if there was insufficient data (i.e., no matching items available or the key equipment, commodity, diagnostic or human resource item required to deliver the intervention was not available) or if the intervention was combined with another intervention due to overlap of the content of care. We aimed to develop QoC indices that reflected recommended interventions based on latest WHO guidelines for the three subpopulations (women who recently delivered, newborns, SSNs) and the items required to deliver those interventions. As such, all interventions that met the inclusion criteria and all items within those interventions that were an exact match, high partial match, or low partial match were included in the QoC indices. We reviewed the exact and partial match items across interventions and identified interventions for which items overlapped.

Interventions for which all matching items were also included in another intervention were combined. We also examined the balance of items across interventions and combined some items into a single indicator (e.g., immunization supplies, available HIV guidelines and staff training, training in integrated management of pregnancy and childbirth (IMPAC) or newborn care) to ensure the indices were not dominated by any one intervention – see **Table S3** and **Table S5** in the Online Supplementary Document for detailed indicator definitions.

We assessed possible methods for combining the index items including simple average, weighted average with weighting either by intervention or QoC sub-domain, and data-driven approaches including principal component analysis (PCA), latent class analysis, and item response theory. We ultimately excluded data-driven approaches as they often resulted in indices that did not reflect conceptual frameworks and clinical knowledge of QoC. We considered the distribution of items within QoC sub-domains and within interventions to decide on a simple or weighted average approach to calculating the index scores for each service area. If the number of items was similar in each sub-domain, we opted for a simple average. Otherwise, a weighted average was used with the option to utilize a sub-domain weighted approach or an intervention-weighted approach. Finally, where an intervention-weighted approach was used, we reviewed items across interventions to identify general items that were required for multiple interventions. These items were moved to a separate general intervention area to prevent double counting of items across interventions within the index.

## RESULTS

### Identification of interventions, guideline review and item identification

For maternal PNC, a total of 36 interventions were identified and 21 met the inclusion criteria for guideline review and item identification. Reasons for intervention exclusion included WHO non-recommended interventions (n=6) and non-clinical interventions (n=9). For newborn PNC, a total of 14 interventions were identified and 13 met the inclusion criteria for guideline review and item identification. Reasons for intervention exclusion included non-recommended intervention (n=1). For SSNC, a total of 30 interventions were identified and 19 met the inclusion criteria for guideline review and item identification. Reasons for intervention exclusion included intensive-care level newborn care interventions (n=11) (**Figure 1**). The full list of interventions is available in **Table S1** in the Online Supplementary Document.

**Figure 1.** Intervention selection for maternal PNC, newborn PNC, and SSNC.

**Figure 2.**
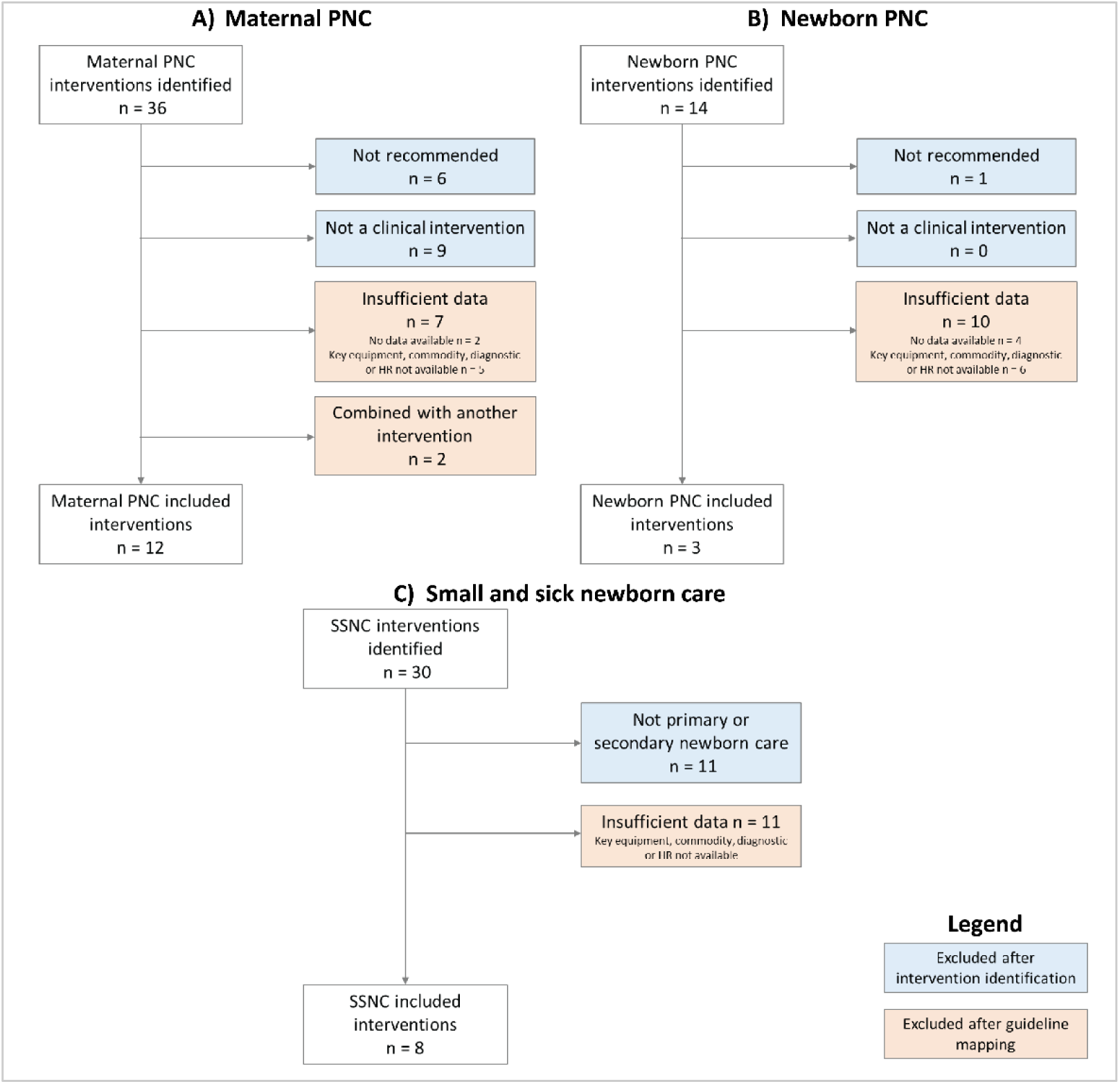
Intervention selection for maternal PNC, newborn PNC, and SSNC

Guideline extraction was conducted for the 21 maternal PNC interventions, 13 newborn PNC interventions, and 19 SSNC interventions. Notably, there was variability in the total number of items required for each intervention as well as the sub-domains across which those items were located. A detailed list of guideline extraction items is in **Table S2** in the Online Supplementary Document.

### Data mapping

#### Overview of mapping and quality of alignments

Review of the SPA and SARA revealed that provision and experience of care data collected through direct observation and client exit interviews was limited to a few select services (antenatal care, family planning, curative care for sick children). While the updated 2023 SPA contains an exit interview for PNC clients, it only covers topics related to counseling and experience of care, which is insufficient to develop a full provision/experience of care index. While there is evidence of validity of maternal report of certain PNC interventions through exit interviews, those interventions were largely not included in the SPA exit interview [32,33]. The lack of provision and experience of care data resulted in no matching items for these quality domains.

**Table 1**, **Table 2**, and **Table 3** show the results from mapping extracted readiness items to HFA questionnaires. For maternal PNC, across the 21 included maternal interventions from the WHO PNC guidelines [8], there were 89 full or partial matches and 40 nonmatches; for newborn PNC, across the 13 included newborn interventions from the WHO PNC guidelines [8], there were 44 full or partial matches and 48 nonmatches, and for SSNC, across the 19 included from the review of WHO guidelines and previous studies assessing facility readiness for SSNC [7,24–26], there were 167 full or partial matches and 130 nonmatches.

**Table 1.**
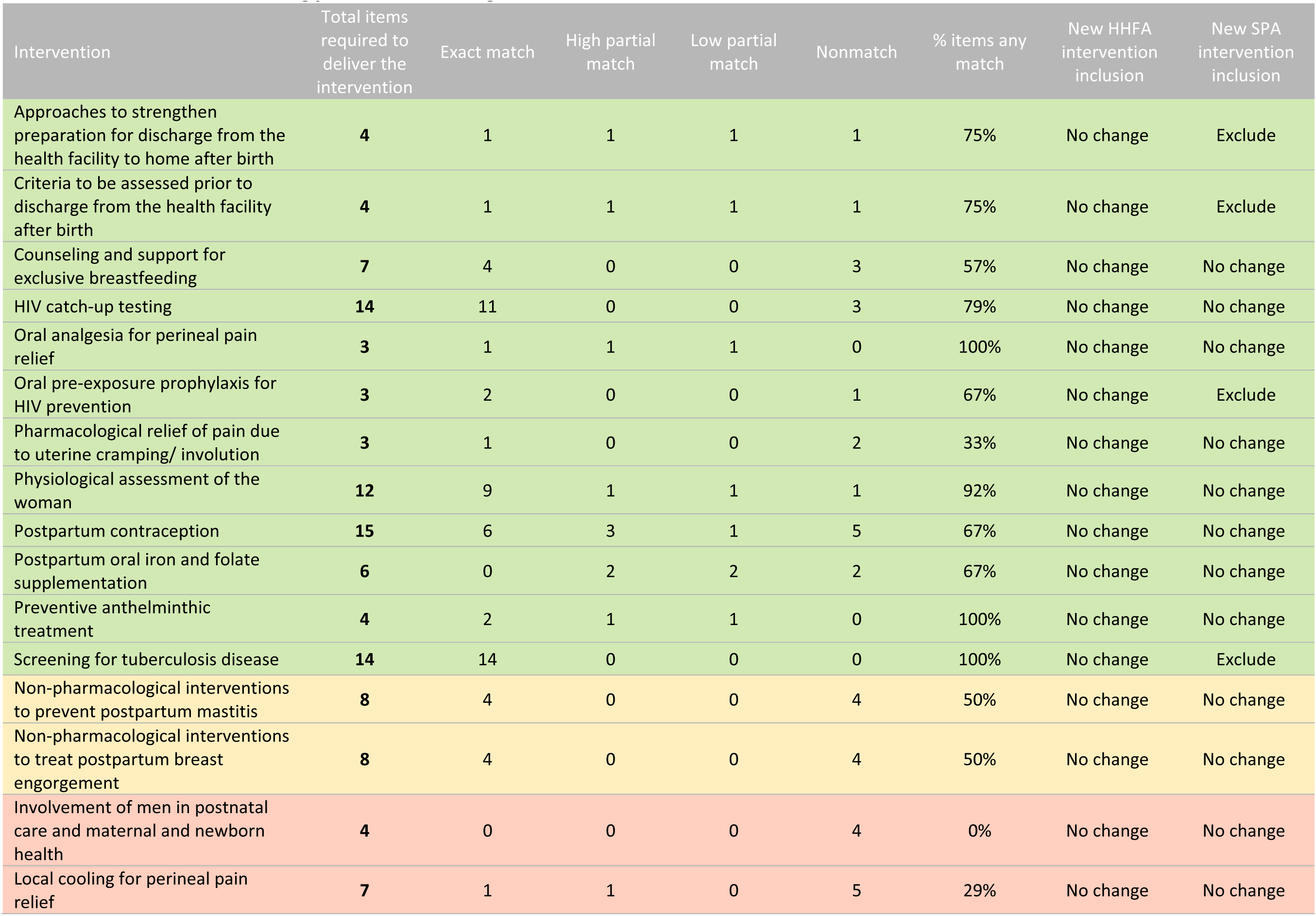
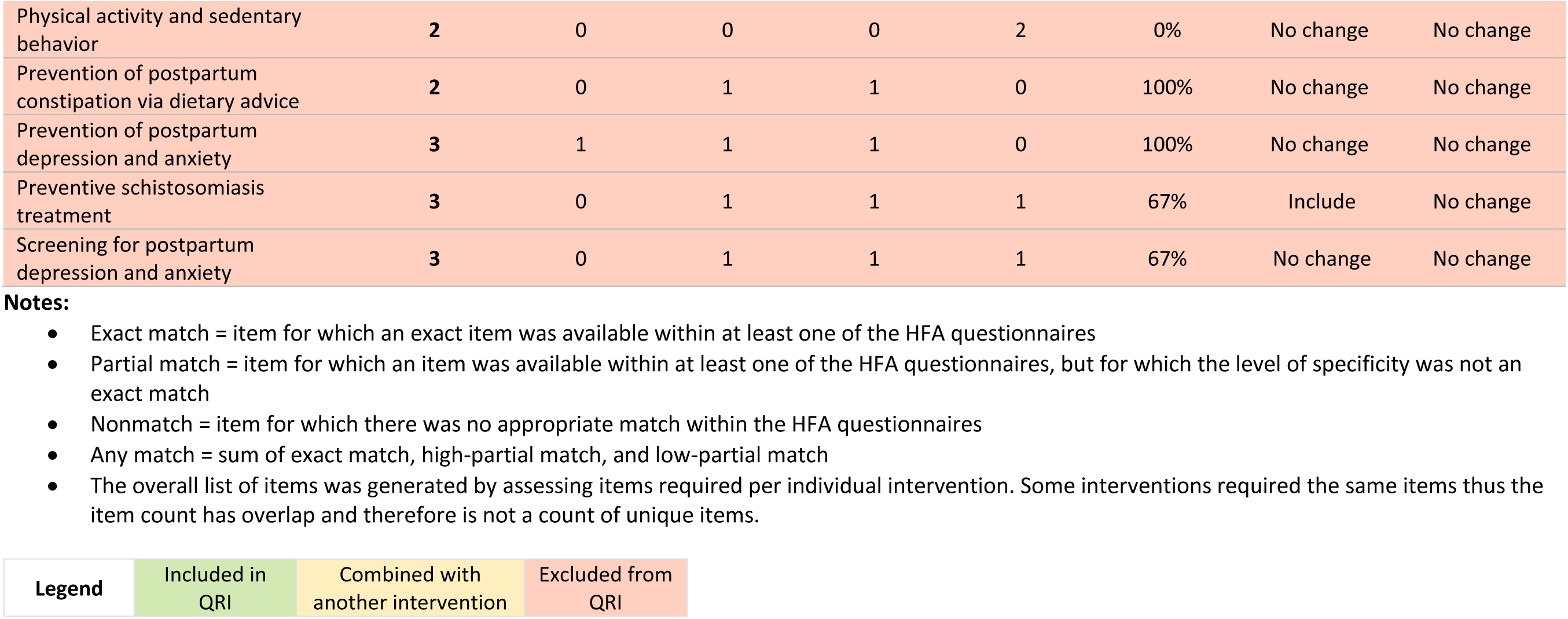
Readiness item matching from intervention guidelines to HFA items, maternal PNC.

| Intervention | Total items required to deliver the intervention | Exact match | High partial match | Low partial match | Nonmatch | % items any match | New HHFA intervention inclusion | New SPA intervention inclusion |
| --- | --- | --- | --- | --- | --- | --- | --- | --- |
| Approaches to strengthen preparation for discharge from the health facility to home after birth | 4 | 1 | 1 | 1 | 1 | 75% | No change | Exclude |
| Criteria to be assessed prior to discharge from the health facility after birth | 4 | 1 | 1 | 1 | 1 | 75% | No change | Exclude |
| Counseling and support for exclusive breastfeeding | 7 | 4 | 0 | 0 | 3 | 57% | No change | No change |
| HIV catch-up testing | 14 | 11 | 0 | 0 | 3 | 79% | No change | No change |
| Oral analgesia for perineal pain relief | 3 | 1 | 1 | 1 | 0 | 100% | No change | No change |
| Oral pre-exposure prophylaxis for HIV prevention | 3 | 2 | 0 | 0 | 1 | 67% | No change | Exclude |
| Pharmacological relief of pain due to uterine cramping/ involution | 3 | 1 | 0 | 0 | 2 | 33% | No change | No change |
| Physiological assessment of the woman | 12 | 9 | 1 | 1 | 1 | 92% | No change | No change |
| Postpartum contraception | 15 | 6 | 3 | 1 | 5 | 67% | No change | No change |
| Postpartum oral iron and folate supplementation | 6 | 0 | 2 | 2 | 2 | 67% | No change | No change |
| Preventive anthelmintic treatment | 4 | 2 | 1 | 1 | 0 | 100% | No change | No change |
| Screening for tuberculosis disease | 14 | 14 | 0 | 0 | 0 | 100% | No change | Exclude |
| Non-pharmacological interventions to prevent postpartum mastitis | 8 | 4 | 0 | 0 | 4 | 50% | No change | No change |
| Non-pharmacological interventions to treat postpartum breast engorgement | 8 | 4 | 0 | 0 | 4 | 50% | No change | No change |
| Involvement of men in postnatal care and maternal and newborn health | 4 | 0 | 0 | 0 | 4 | 0% | No change | No change |
| Local cooling for perineal pain relief | 7 | 1 | 1 | 0 | 5 | 29% | No change | No change |
| Physical activity and sedentary behavior | 2 | 0 | 0 | 0 | 2 | 0% | No change | No change |
| Prevention of postpartum constipation via dietary advice | 2 | 0 | 1 | 1 | 0 | 100% | No change | No change |
| Prevention of postpartum depression and anxiety | 3 | 1 | 1 | 1 | 0 | 100% | No change | No change |
| Preventive schistosomiasis treatment | 3 | 0 | 1 | 1 | 1 | 67% | Include | No change |
| Screening for postpartum depression and anxiety | 3 | 0 | 1 | 1 | 1 | 67% | No change | No change |

**Notes:**

|  |  |  |  |
| --- | --- | --- | --- |
| **Legend** | Included in QRI | Combined with another intervention | Excluded from QRI |

**Table 2.**
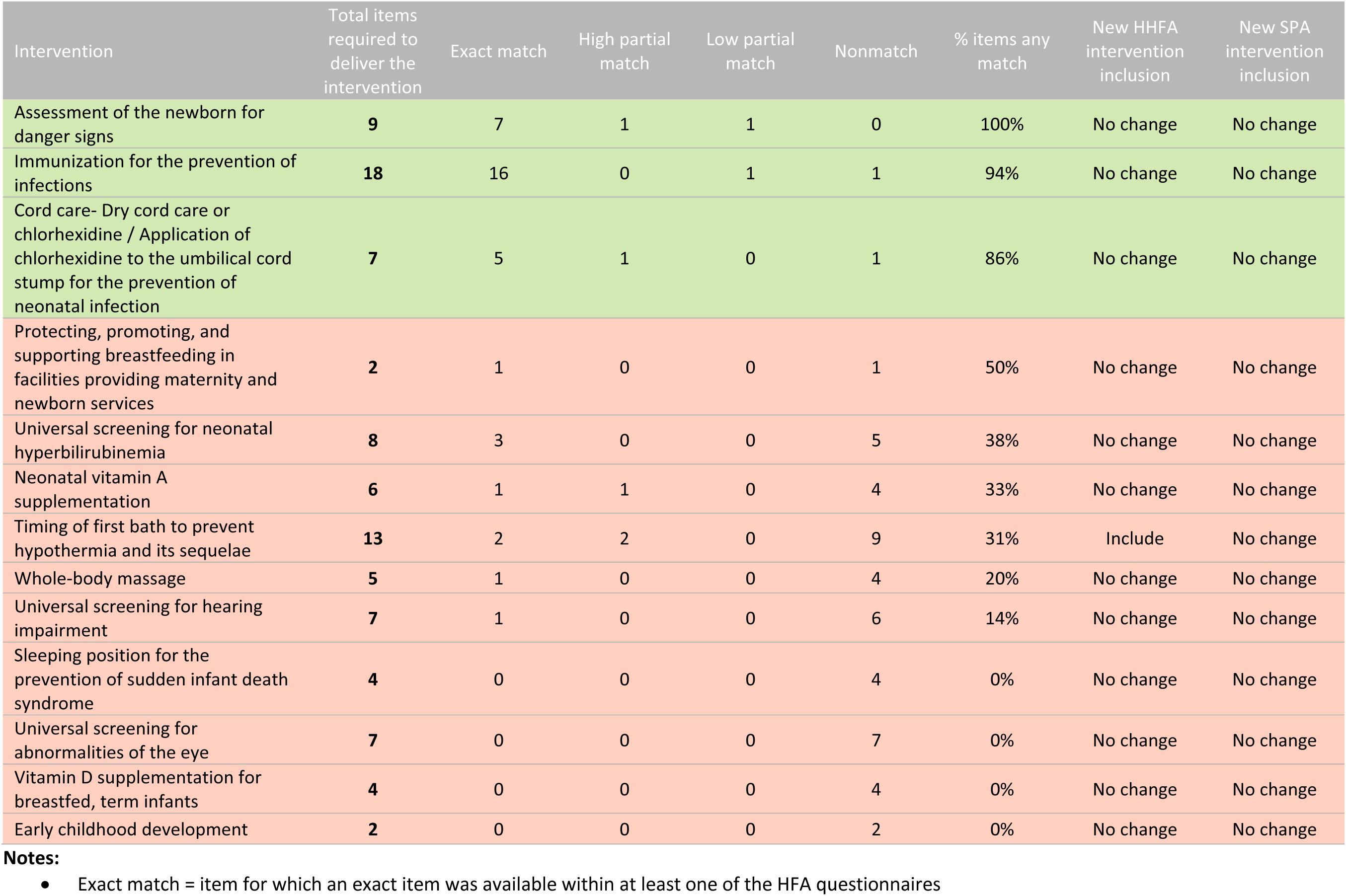
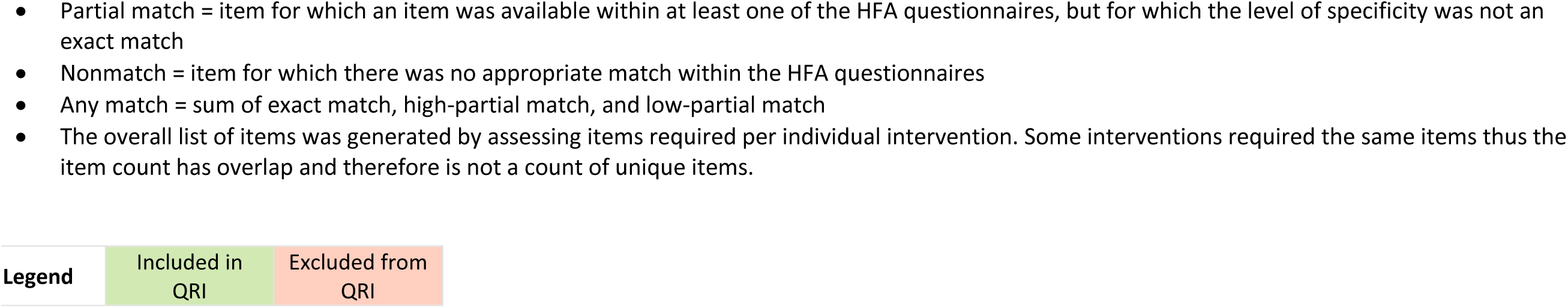
Readiness item matching from intervention guidelines to HFA items, newborn PNC.

| Intervention | Total items required to deliver the intervention | Exact match | High partial match | Low partial match | Nonmatch | % items any match | New HHFA intervention inclusion | New SPA intervention inclusion |
| --- | --- | --- | --- | --- | --- | --- | --- | --- |
| Assessment of the newborn for danger signs | 9 | 7 | 1 | 1 | 0 | 100% | No change | No change |
| Immunization for the prevention of infections | 18 | 16 | 0 | 1 | 1 | 94% | No change | No change |
| Cord care- Dry cord care or chlorhexidine / Application of chlorhexidine to the umbilical cord stump for the prevention of neonatal infection | 7 | 5 | 1 | 0 | 1 | 86% | No change | No change |
| Protecting, promoting, and supporting breastfeeding in facilities providing maternity and newborn services | 2 | 1 | 0 | 0 | 1 | 50% | No change | No change |
| Universal screening for neonatal hyperbilirubinemia | 8 | 3 | 0 | 0 | 5 | 38% | No change | No change |
| Neonatal vitamin A supplementation | 6 | 1 | 1 | 0 | 4 | 33% | No change | No change |
| Timing of first bath to prevent hypothermia and its sequelae | 13 | 2 | 2 | 0 | 9 | 31% | Include | No change |
| Whole-body massage | 5 | 1 | 0 | 0 | 4 | 20% | No change | No change |
| Universal screening for hearing impairment | 7 | 1 | 0 | 0 | 6 | 14% | No change | No change |
| Sleeping position for the prevention of sudden infant death syndrome | 4 | 0 | 0 | 0 | 4 | 0% | No change | No change |
| Universal screening for abnormalities of the eye | 7 | 0 | 0 | 0 | 7 | 0% | No change | No change |
| Vitamin D supplementation for breastfed, term infants | 4 | 0 | 0 | 0 | 4 | 0% | No change | No change |
| Early childhood development | 2 | 0 | 0 | 0 | 2 | 0% | No change | No change |

**Notes:**

|  |  |  |
| --- | --- | --- |
| **Legend** | Included in<br>QRI | Excluded from<br>QRI |

**Table 3.** Readiness item matching from intervention guidelines to HFA items, small and/or sick newborn care.

| Intervention | Total items required to deliver the intervention | Exact match | High partial match | Low partial match | Nonmatch | % items any match | New HHFA intervention inclusion | New SPA intervention inclusion | Overlap with PNC interventions |
| --- | --- | --- | --- | --- | --- | --- | --- | --- | --- |
| Prevention of mother-to-child transmission of HIV | 19 | 16 | 0 | 0 | 3 | 84% | No change | Exclude | Yes |
| Immediate newborn care and Routine care | 33 | 24 | 1 | 0 | 8 | 76% | No change | No change | Yes |
| Detection and management of neonatal infection (injectable antibiotics) | 22 | 15 | 0 | 0 | 7 | 68% | No change | No change | No |
| Comfort and pain management | 12 | 8 | 0 | 0 | 4 | 67% | No change | No change | No |
| Early initiation and support for breastfeeding | 6 | 4 | 0 | 0 | 2 | 67% | No change | No change | Yes |
| Detection and management of hypoglycemia | 14 | 7 | 1 | 0 | 6 | 57% | No change | No change | No |
| Neonatal resuscitation | 20 | 11 | 0 | 0 | 9 | 55% | No change | No change | No |
| Kangaroo mother care | 17 | 6 | 0 | 0 | 11 | 35% | No change | Exclude | No |
| Prevention of apnea | 9 | 6 | 0 | 0 | 3 | 67% | Include | No change | No |
| Seizure management | 9 | 6 | 0 | 0 | 3 | 67% | Include | Include | No |
| Detection and management of anemia | 27 | 17 | 0 | 0 | 10 | 63% | No change | Include | No |
| Detection and management of neonatal encephalopathy | 7 | 4 | 0 | 0 | 3 | 57% | No change | No change | No |
| Detection and referral management of birth defects | 2 | 1 | 0 | 0 | 1 | 50% | No change | No change | No |
| Pre-discharge advice on mother and baby care and follow up | 6 | 3 | 0 | 0 | 3 | 50% | No change | No change | Yes |
| Safe administration of intravenous fluids | 21 | 10 | 0 | 0 | 11 | 48% | No change | No change | No |
| Detection and management of jaundice (phototherapy) | 26 | 11 | 0 | 0 | 15 | 42% | No change | No change | Yes |
| Thermal care | 12 | 5 | 0 | 0 | 7 | 42% | Include | No change | No |
| Safe administration of oxygen | 15 | 5 | 0 | 0 | 10 | 33% | Include | Include | No |
| Assisted feeding for optimal nutrition | 20 | 5 | 1 | 0 | 14 | 30% | No change | No change | No |

**Notes:**

| Legend | Included in QRI | Excluded from QRI |
| --- | --- | --- |

Many of the maternal and newborn PNC exact matches were service readiness items required to provide health services generally, but were not specific to PNC (e.g., infection prevention and control items, power, vaccines, diagnostics for tuberculosis (TB) and human immunodeficiency virus (HIV), thermometer, stethoscope). Partial and nonmatches reflected common limitations in the SPA and SARA. For example, there was a lack of items on specific PNC training topics and a lack of specificity about guideline content for PNC services. No dosage information was captured in SPA and SARA for medicines/ commodities, although this is required to determine readiness for interventions by age group (e.g., neonatal and maternal vitamin A supplementation, maternal and neonatal iron supplementation). In addition, some PNC-specific commodities were not collected at all (e.g., vitamin D, massage oil). PNC-specific equipment and supplies such as materials required to conduct universal screenings (e.g., for hearing, eye abnormalities, neonatal hyperbilirubinemia) and education materials were not captured in the SPA and SARA questionnaires.

For SSNC, exact matches included service readiness items required to provide health services generally (e.g., power, emergency transportation, hemoglobin testing, full blood count, antiretrovirals, vaccines) as well as some SSNC-specific items (e.g., neonatal bag and mask device, weighing scale, first line antibiotics). Partial and nonmatches reflected a lack of special newborn care clinical interventions for SSNC in the SPA and SARA questionnaires. In addition, some SSNC-specific commodities were not collected at all (e.g., vitamin K, oral sucrose, phenobarbital, methylxanthines). SSNC-specific equipment and supplies were also limited in scope in the SPA and SARA questionnaires. In addition, like PNC, there was a lack of items on specific SSNC training topics and a lack of specificity regarding guideline content for SSNC services.

#### Intervention inclusion and exclusion based on data availability

For maternal PNC, twelve interventions were retained for inclusion in the maternal PNC quality readiness index (QRI) (**Table 1**, interventions highlighted in green). There were seven interventions with insufficient data for inclusion in the maternal PNC-QRI (**Figure 1**). Two interventions had no matching items in the HFAs, and five interventions were missing the key equipment, commodity, diagnostic or human resources required to deliver the intervention. Interventions that were excluded based on data availability were often counselling-based or required only trained staff. In addition, two interventions (non-pharmacological interventions to prevent breast engorgement and postpartum mastitis) were excluded as they were combined with another intervention (counseling and support for exclusive breastfeeding) due to overlap in the content of care and required readiness items with available data.

For newborn PNC, three interventions were retained for inclusion in the newborn PNC-QRI (**Table 2**, interventions highlighted in green). There were ten interventions with insufficient data for inclusion in the newborn PNC-QRI (**Figure 1**). Four interventions had no matching items in the HFAs, and six interventions were missing the key equipment, commodity, diagnostic or human resources required to deliver the intervention. Interventions that were excluded based on data availability were often universal screenings that required specific equipment or interventions requiring specific commodities that were not available in the HFAs.

For SSNC, eight interventions were retained for inclusion in the SSNC-QRI (**Table 3**, interventions highlighted in green). We found that there was an overlap between newborn PNC and SSNC interventions. This is likely because many small and/or sick newborns also need essential PNC services (PMTCT, immediate newborn care, early initiation and support for breastfeeding, pre-discharge advice on mother and baby care and follow up, detection and management of jaundice). There were eleven interventions with insufficient data for inclusion in the SSNC-QRI (**Figure 1**). Interventions that were excluded were often services expected to be available at hospitals with specialized newborn care units. All these interventions were missing the key equipment, commodity, diagnostic or HR required to deliver the intervention.

#### Mapping and quality of alignment with new SPA and HHFA

Both SPA and SARA surveys had a few notable limitations for measuring the quality of PNC and SSNC services. Neither facility survey included a specific service area module for collecting data on PNC or SSNC. Instead, these instruments relied on modules covering other service areas where PNC and SSNC services may be delivered (e.g., childbirth, child well-visits, HIV/AIDS) to gather information about readiness to deliver PNC and SSNC. In addition, the SPA and SARA do not collect any provision and experience of care items for PNC and SSNC. However, both surveys have recently (in 2022) undergone a revision process and there is a new SPA questionnaire and the HHFA questionnaire to replace the SARA [21,31]. We repeated the mapping exercise using the new versions of the SPA and HHFA questionnaires (**Table 1**, **Table 2**, and **Table 3**). We found that in general, the HHFA expanded to include a specific section for PNC and SSNC along with additional medicines, equipment, and supplies for interventions such as thermal care, and inclusion of more specific PNC guidelines and staff training. The SPA, in comparison, largely contracted with a reduction of items, to be a more streamlined tool with no specific PNC or SSNC sections. These changes mean that for the maternal PNC-QRI, the HHFA would allow for inclusion of one additional context-specific intervention (preventive schistosomiasis treatment) while the SPA would result in exclusion of four additional interventions, due to items being dropped from the new survey version (**Table 1**). For the newborn PNC-QRI, the only change if using the HHFA survey would be the ability to include timing of first bath to prevent hypothermia and its sequelae (**Table 2**). Mapping to the new SPA and HHFA questionnaires resulted in more changes for the SSNC-QRI. The HHFA would allow for inclusion of four additional interventions while the SPA would allow for the inclusion of three additional interventions, two of which are the same as the HHFA. However, the new SPA would also result in the exclusion of two interventions, PMTCT and KMC (**Table 3**).

#### Final index development

No provision/experience of care data was available for PNC or SSNC, thus the indices reflect facility readiness only. After exclusion of interventions with insufficient data, limited data was available to generate readiness indices for maternal PNC, newborn PNC, and SSNC. As a result, we did not conduct an expert survey to prioritize interventions or items within interventions as has been done for other service areas [23,34,35]. Instead, we included all available items based on the older SPA and SARA survey mapping for each included intervention in the indices. We also examined the balance of items across interventions and combined some items into a single indicator to ensure the indices were not dominated by any one intervention (e.g., immunization supplies, available HIV guidelines and staff training, training in IMPAC or newborn care – see **Table S3**, **Table S4**, and **Table S5** in the Online Supplementary Document for detailed indicator definitions). We retained and denoted context-specific interventions and associated items in the QRIs so that their inclusion can operationalized based on country policy. For the maternal PNC-QRI and newborn PNC-QRI, there was an unequal distribution of items across sub-domains and similar individual items across interventions; thus, we opted for a sub-domain weighted approach with the sub-domains corresponding to basic amenities, equipment and supplies, medicines and commodities, diagnostics, and guidelines and staff training. For the SSNC-QRI, in contrast, there was an unequal distribution of items across sub-domains and across interventions; thus, we opted for the intervention weighted approach. It is also important to note that interventions needed for small newborns are sometimes different from the interventions needed for sick newborns. We have included all interventions together in this index, but it may be important to look at the quality of individual interventions if interested in assessing readiness for small newborns separately from sick newborns.

The proposed items for QRIs for maternal PNC, newborn PNC, and SSNC are in **Table 4**, **Table 5**, and **Table 6**. The maternal PNC-QRI includes 24 items, of which eight items are context specific, across 5 sub-domains– 11 equipment and supplies (of which five are infection prevention and control related items), five medicines and commodities, two diagnostics, one basic amenity, and five guidelines and staff training. The newborn PNC-QRI includes 16 items, of which one is context specific, across three sub-domains – 11 equipment and supplies (of which five are infection prevention and control related items), two medicines and commodities, and three guidelines and staff training. The SSNC-QRI includes 48 items, of which nine are context specific, across eight interventions plus an intervention for general/cross cutting readiness items. The interventions with the most items are immediate newborn care and routine care (11 items), PMTCT (eight items), and general readiness items (12 items) while the interventions with the fewest items are early initiation and support for breastfeeding (two items), comfort and pain management (two items), and detection and management of hypoglycemia (two items).

**Table 4.**
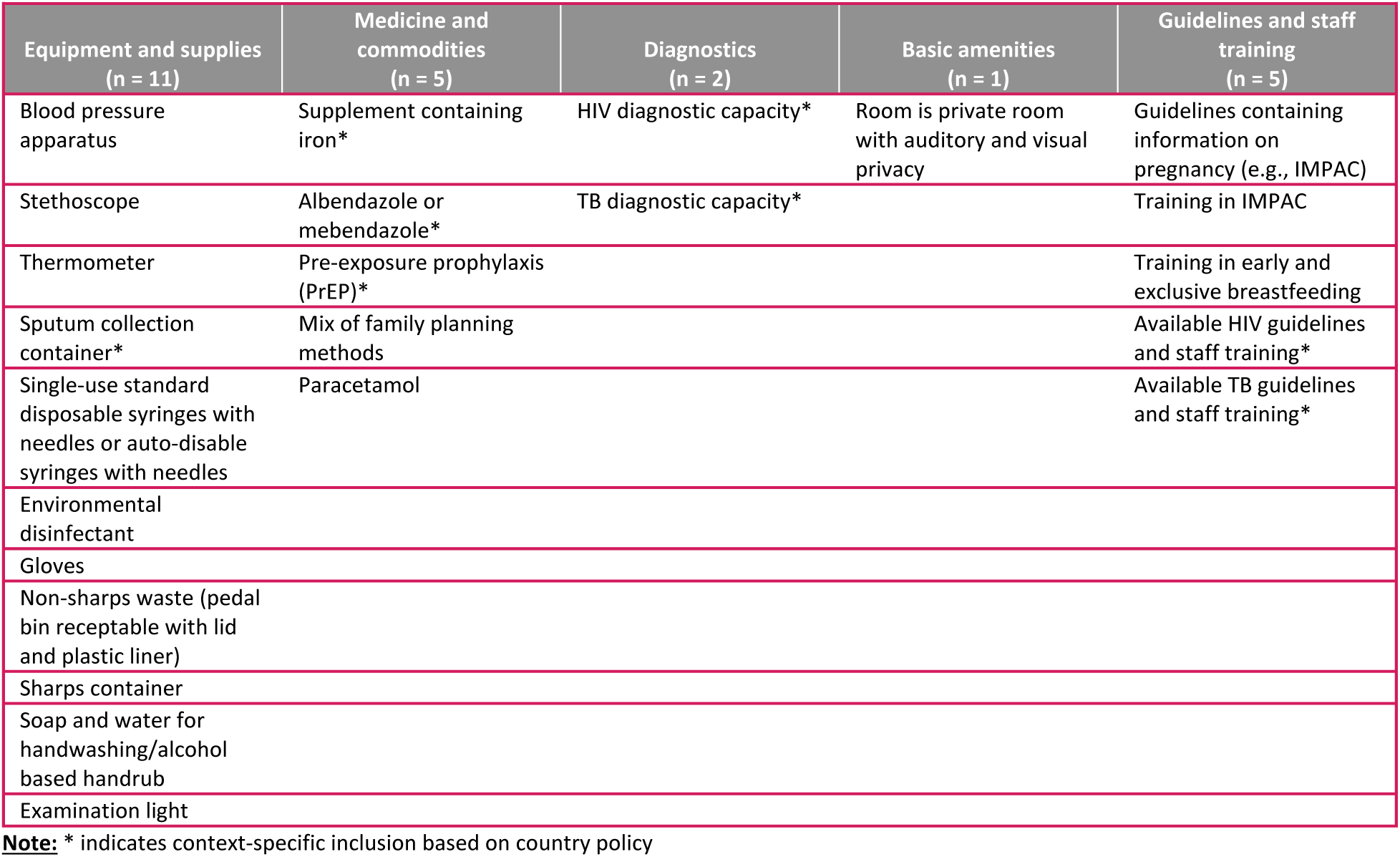
Proposed items in the maternal PNC-QRI by readiness sub-domains.

| Equipment and supplies<br>(n = 11) | Medicine and commodities<br>(n = 5) | Diagnostics<br>(n = 2) | Basic amenities<br>(n = 1) | Guidelines and staff training<br>(n = 5) |
| --- | --- | --- | --- | --- |
| Blood pressure apparatus | Supplement containing iron* | HIV diagnostic capacity* | Room is private room with auditory and visual privacy | Guidelines containing information on pregnancy (e.g., IMPAC) |
| Stethoscope | Albendazole or mebendazole* | TB diagnostic capacity* |  | Training in IMPAC |
| Thermometer | Pre-exposure prophylaxis (PrEP)* |  |  | Training in early and exclusive breastfeeding |
| Sputum collection container* | Mix of family planning methods |  |  | Available HIV guidelines and staff training* |
| Single-use standard disposable syringes with needles or auto-disable syringes with needles | Paracetamol |  |  | Available TB guidelines and staff training* |
| Environmental disinfectant |  |  |  |  |
| Gloves |  |  |  |  |
| Non-sharps waste (pedal bin receptable with lid and plastic liner) |  |  |  |  |
| Sharps container |  |  |  |  |
| Soap and water for handwashing/alcohol based handrub |  |  |  |  |
| Examination light |  |  |  |  |

**Note:** \* indicates context-specific inclusion based on country policy

**Table 5.** Proposed items in the newborn PNC-QRI by readiness sub-domains.

| Equipment and supplies<br>(n = 11) | Medicine and commodities<br>(n = 2) | Diagnostics<br>(n = 0) | Basic amenities<br>(n = 0) | Guidelines and staff training<br>(n = 3) |
| --- | --- | --- | --- | --- |
| Stethoscope | Chlorhexidine solution* |  |  | IMPAC or ENC guidelines |
| Thermometer | Vaccines (BCG, Hep B, OPV0) |  |  | Child vaccination guidelines |
| Single-use standard disposable syringes with needles or auto-disable syringes with needles |  |  |  | Training in IMPAC or newborn care |
| Vaccine documentation (Blank/unused individual child vaccination cards or booklets, and immunization tally sheet) |  |  |  |  |
| Refrigerator with temperature monitoring device and power; or vaccine carrier with ice packs |  |  |  |  |
| Gloves |  |  |  |  |
| Environmental disinfectant |  |  |  |  |
| Non-sharps waste (pedal bin receptable with lid and plastic liner) |  |  |  |  |
| Room with auditory and visual privacy |  |  |  |  |
| Soap and water for handwashing/alcohol based handrub |  |  |  |  |
| Sharps container |  |  |  |  |

**Note:** \* indicates context-specific inclusion based on country policy

**Table 6.**
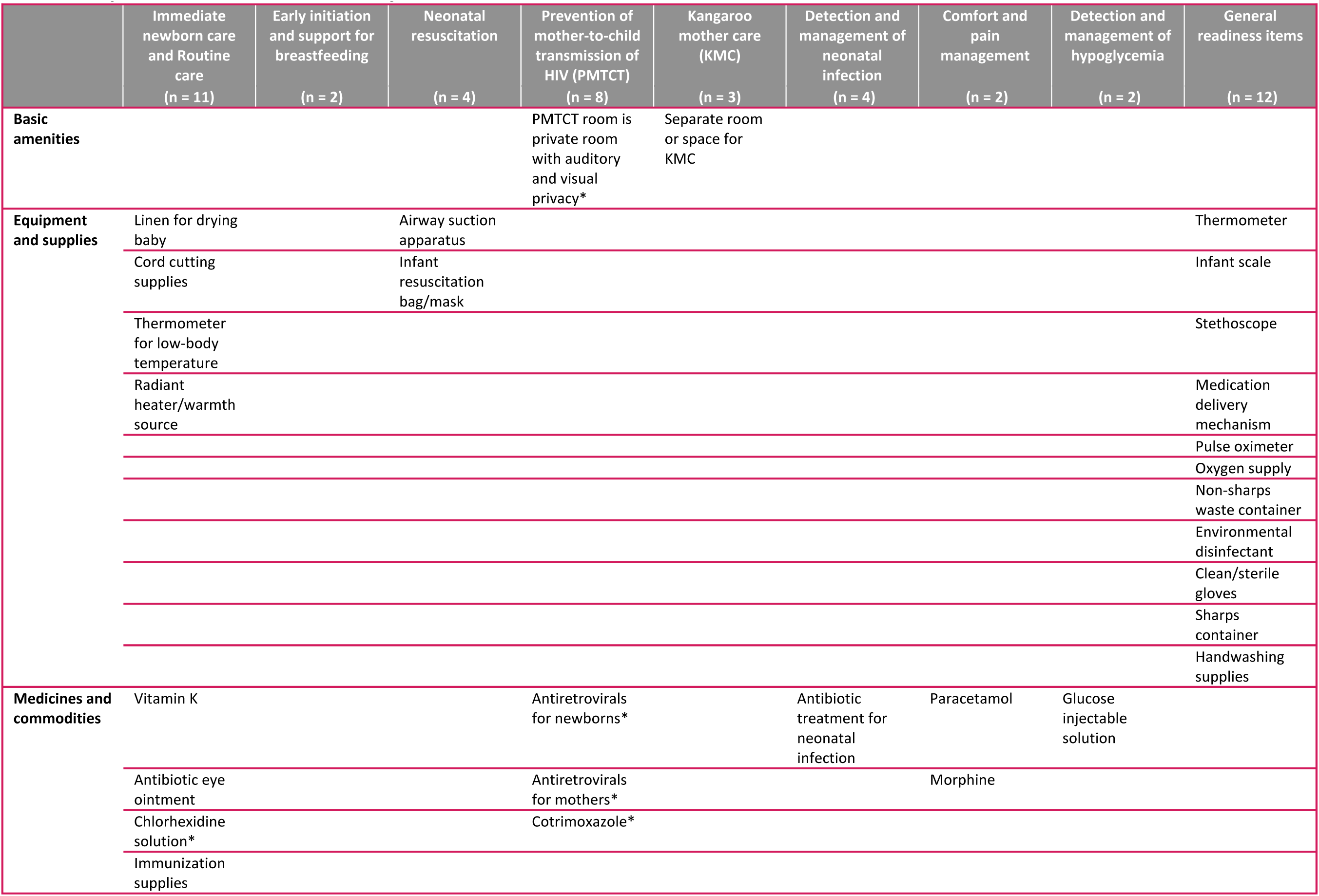
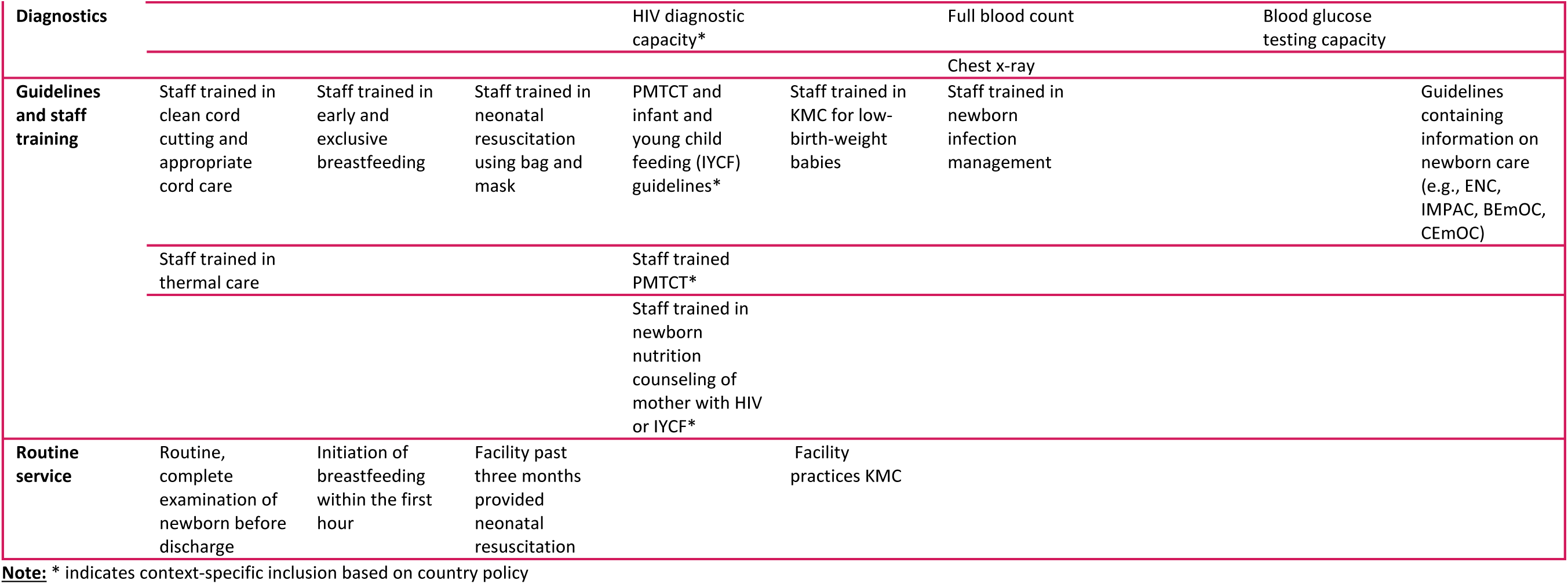
Proposed items in the SSNC-QRI by intervention and readiness sub-domains.

## DISCUSSION

We describe the development of three QRIs that can provide standardized measures for maternal PNC, newborn PNC, and SSNC service readiness and can be adapted at country level and operationalized using existing HFA data, facilitating their use by decision-makers for planning and resource allocation. A lack of data currently availability in HFAs meant that we could not develop summary indices of service provision or experience of care. There were also substantial gaps in the readiness data even after reviewing the recently revised SPA and HHFA.

There have been a few attempts to systematically develop indices of quality of care for maternal and newborn health services that have carefully described their methods and assumptions which have focused on family planning, antenatal care, nutrition, and childbirth care [23,34,36,37]. These approaches are similar to our approach described in this manuscript for developing summary QRIs for maternal PNC, newborn PNC, and SSNC in that they utilize a systematic approach to index development, are rooted in latest guidelines and guidance, and explore multiple approaches to item selection and index aggregation methods. Much more commonly, research studies exploring access to quality services and/or associations between quality and other outcomes (for example many of the studies included in the Do et al and Sheffel et al reviews [28,38]) use summary measures of service quality, but the methods for the development of these summary measures are secondary to the primary research question. As a result, there is substantial variability in the methods employed including item inclusion and aggregation methods, largely due to the lack of guidance on best practices. Thus, it is difficult to synthesize learnings across these studies, as they may not be comparable due to inconsistencies in measurement approaches. Our work on developing summary QRIs for maternal PNC, newborn PNC, and SSNC utilizing a systematic approach to identifying interventions and items and guided by up-to-date clinical guidelines contributes to the growing evidence around generating summary measures of service quality.

The data mapping process, which assessed data gaps across two versions of two different HFAs (the SARA/HHFA and the SPA), highlighted limitations of existing HFAs to characterize service readiness for PNC and SSNC. This finding echoes other studies which have noted the need to align existing measurement tools with global standards in order to fill gaps in quality of care measurement [28,29]. We found that publicly available HFAs have not historically included a PNC or SSNC module hence have had very limited readiness data for the target groups. However, the new HHFA does have PNC and SSNC modules and additional PNC and SSNC readiness items have been added to the HHFA, which will be beneficial for assessing service readiness moving forward. We also found that there are some recommended interventions completely omitted from HFAs. Our SSNC-QRI could not include any intensive level interventions, as HFAs have primarily been designed to collect information at primary/secondary level health care facilities. The PNC interventions we excluded due to data insufficiency were often counselling-based and required only trained staff, or universal screening (e.g. hearing or eye abnormality) that required specific equipment. We were also limited to developing only service readiness indices as there is no direct observation of PNC or SSNC in the SPA/SARA/HHFA to enable measurement of process quality. Finally, many of the exact matches found in the existing HFAs were service readiness items required to provide health services generally and were not specific to PNC and SSNC. This finding may have implications for the ability of these indices to differentiate facilities with high and low readiness for PNC and SSNC. Research has shown that indices generated with relatively few items are prone to ties across facilities and ceiling effects, particularly when many of the items are almost universally available at health facilities [37]. In the absence of a validated set of tracer indicators, efforts to strengthen the comprehensive measurement of maternal PNC, newborn PNC, and SSNC readiness are warranted, with a focus on including items in HFAs based on clinical considerations and the ability to discriminate between levels of service readiness.

The main advantage of a summary measure is to allow monitoring of progress and comparisons of the levels and trends in service readiness for maternal and newborn care at national level [39]. Although composite measures like QRIs do not provide information on which specific items are lagging behind, this information can readily be obtained by policymakers and stakeholders at a national and sub-national level if needed to inform targeted interventions and resource allocation. Summary measures of service readiness may also be useful for conducting effective coverage analyses that examine readiness (i.e. input-adjusted coverage) as a key step in the effective coverage cascade [40]. However, summary indices may be less useful at the facility level, where more granular information may be required to identify specific problem areas for quality improvement. Our proposed maternal PNC, newborn PNC, and SSNC QRIs will require country adaptation, especially for context-specific interventions, which may make cross-country comparisons more difficult at regional or global levels.

The study has several limitations that should be acknowledged. First, the reliance on existing data sources limited the scope of the indices, as some interventions and domains of quality could not be adequately captured. One key challenge with the SSNC-QRI is the exclusion of intensive/transition interventions (e.g. CPAP) from the readiness index. These interventions are linked to ENAP coverage measures; however, these interventions could not be included in our QRI due to data availability gaps. Although most small and/or sick newborns do not require intensive care, expansion of HFAs to capture these interventions may be helpful in filling this data gap. It may also be difficult to incorporate these intensive-level interventions into a QRI, as it will require facilities at distinct levels to have different indices to account for differentials in expected service delivery. Hence using existing HFA data to improve measurement for intervention readiness that will benefit the greatest number of SSNs can be prioritized now and intensive level interventions added into further versions of these HFA tools. The current data availability gaps for provision and experience of care also limited our ability to assess the technical delivery of PNC and SSNC interventions and the experiences of mothers and newborns. As a result, the indices we developed represent a set of items that can be measured with existing HFA data to facilitate country use of these measures given current data constraints. However, they are not representative of readiness to deliver complete maternal/ newborn PNC or SSNC services. Second, we did not conduct an expert survey to prioritize readiness items for inclusion in the indices which would strengthen the face validity of the indices. There were few items available in existing HFAs thus, there was no need to prioritize the available items. However, we utilized a guideline-driven approach to item selection, prioritizing recently published service guidelines such as the WHO PNC guidelines [8], Standards for improving the quality of care for small and sick newborns in health facilities [7], and the WHO framework for the provision of quality maternal and newborn care [9], which were developed through an extensive literature review and expert consultations. Third, while the indices are designed to measure what should be happening in health service delivery, their generalizability may vary across countries due to differences in health systems and implementation of interventions, which can affect the applicability of the indices. While we have proposed a single index based on guidelines and data availability, adaptation and validation of the indices at the country level is necessary to ensure they are suitable to the setting in which they are used. Finally, we were not able to assess construct validity by, for example, examining the association with provision of care or health outcomes.

Our work has highlighted a number of critical areas for future research. Future efforts to develop an ideal summary measure of service readiness and provision/experience of care for maternal PNC, newborn PNC, and SSNC without consideration of data availability would be helpful to clearly identify data gaps. In addition, identifying a smaller set of salient interventions that are strongly associated with leading causes of death or complications or a small set of items within interventions that are strongly associated with service quality or health outcomes would be helpful to reduce the overall number of items in the index, and thus the data collection burden, and could potentially inform the weighting of items in the index. A more focused set of interventions and items may facilitate measurement of maternal PNC, newborn PNC, and SSNC service quality in HFAs, which must balance comprehensiveness with implementation feasibility. Finally, exploration of alternative data sources such as routine data to generate summary measures of maternal PNC, newborn PNC, and SSNC service readiness and provision/experience of care would be useful in supporting more regular measurement at the country level.

## CONCLUSION

Use of improved data on service readiness and service provision is needed to improve quality of maternal and newborn care going forward. Our summary indices provide a valuable step towards measuring and monitoring service readiness for maternal PNC, newborn PNC, and SSNC in LMICs. The utilization of existing data sources and a systematic approach to index development enhance the feasibility and applicability of these measures. The indices can inform policy and decision-making processes, allowing for targeted interventions and resource allocation to improve the quality of care received by mothers and newborns. Future research to expand the scope of the indices by incorporating provision of care and experience of care domains will be needed. In addition, assessing the proposed indices for validity and reliability would strengthen their effectiveness in capturing the quality of care provided to mothers and newborns. If the gaps in readiness and provision of care measurement are addressed, PNC and SSNC indices have the potential to drive improvements in the delivery of high-quality health services to women and newborns, ultimately contributing to the reduction of maternal and newborn mortality in LMICs.

### DECLARATIONS

## Data Availability

All data produced in the present work are contained in the manuscript.

## Acknowledgements

The authors wish to acknowledge the Bill & Melinda Gates Foundation for their support of this project. We are grateful to the Improving Measurement & Program Design (IMPROVE) core group for their technical inputs and expertise during the development of this analysis, in particular Allisyn Moran. We additionally would like to thank Amy Hobbs for her contributions to our early work on SSNC readiness indices.

## Ethics approval

This study mapped items in service guidelines and data collection tools but made no use of primary or secondary data. As such, no ethical approval was required.

## Authors’ contributions

AS, MKM, and SK conceptualized the paper and the analysis. AS and SK conducted the analysis. AS prepared the manuscript. All authors critically reviewed and revised the manuscript.

#### Box 1.

**Key definitions of quality dimensions**

- Provision of care refers to the quality of delivery of interventions by providers to clients (i.e., the content of care), which includes following evidence-based practices for routine care and management of complications.
- Experience of care refers to the client’s experience, including effective communication by the care provider about the services provided, client expectations, and client rights; care provided with respect and preservation of dignity; and client access to emotional and social support of their choice.
- Service readiness refers to the capability of health facilities to provide a service of minimum acceptable standards and is measured by the availability of both physical resources and human resources.
- Routine service includes whether the facility reports delivering key interventions. We have included routine service as a sub-domain for SSNC to make up for the lack of data in other readiness sub-domains for SSNC interventions. If more readiness data were available, the routine service sub-domain could be excluded from the index as it reflects historical service delivery rather than service readiness.

#### Box 2.

**Definition of exact, partial, and nonmatch**

- **Exact matches** were items from the guidance documents for which an exact item was available within at least one of the HFA questionnaires.
- **Partial matches** were items for which a partially matching item was available within at least one of the HFA questionnaires. Partial matches were separated based on the specificity of the HFA item compared to the guidance document. For example:

▪ For specific intervention guidelines (e.g., newborn assessment) from the guidance document, a high partial match in the HFA would be broad service areas guidelines that explicitly include that intervention (e.g., guidelines for integrated management of pregnancy and childbirth (IMPAC)).
▪ For the item “staff trained in administering paracetamol within the context of perineal pain relief”, the HFA indicator of staff trained in a broad service area package (e.g., staff trained in IMPAC) was considered a low partial match because it was not clear whether the specific intervention was included in training.
- **Nonmatches** were items for which there was no appropriate match within any HFA questionnaires.

