## Supplemental material for "Advancing maternal and newborn health care measurement: Developing quality of care indices for postnatal and small and/or sick newborn care in low- and middle-income countries"

### SUPPLEMENTARY TABLES

***Table S1: Interventions assessed for index inclusion for maternal PNC, newborn PNC, and small and sick newborn care***

| Intervention | Recommended intervention | Clinical intervention | Level of care | HFA data available |
|---|---|---|---|---|
| **Maternal PNC** | | | | |
| Physiological assessment of the woman | Recommended | Yes | | Yes |
| Prevention of postpartum depression and anxiety | Recommended | Yes | | No |
| Local cooling for perineal pain relief | Recommended | Yes | | No |
| Oral analgesia for perineal pain relief | Recommended | Yes | | Yes |
| Pharmacological relief of pain due to uterine cramping/ involution | Recommended | Yes | | Yes |
| Non-pharmacological interventions to treat postpartum breast engorgement | Recommended | Yes | | Combined with another intervention |
| Non-pharmacological interventions to prevent postpartum mastitis | Recommended | Yes | | Combined with another intervention |
| Prevention of postpartum constipation via dietary advice | Recommended | Yes | | No |
| Screening for postpartum depression and anxiety | Recommended | Yes | | No |
| Physical activity and sedentary behavior | Recommended | Yes | | No |
| Postpartum contraception | Recommended | Yes | | Yes |
| Postpartum oral iron and folate supplementation | Context-specific | Yes | | Yes |
| HIV catch-up testing | Context-specific | Yes | | Yes |
| Screening for tuberculosis disease | Context-specific | Yes | | Yes |
| Preventive anthelminthic treatment | Context-specific | Yes | | Yes |
| Preventive schistosomiasis treatment | Context-specific | Yes | | No |
| Oral pre-exposure prophylaxis for HIV prevention | Context-specific | Yes | | Yes |
| Postpartum vitamin A supplementation | Not recommended | | | |

| Intervention | Recommended intervention | Clinical intervention | Level of care | HFA data available |
|---|---|---|---|---|
| Pharmacological interventions to treat postpartum breast engorgement | Not recommended | | | |
| Pharmacological interventions to prevent postpartum mastitis | Not recommended | | | |
| Postnatal pelvic flood muscle training (PFMT) for pelvic floor strengthening | Not recommended | | | |
| Prevention of maternal peripartum infection after uncomplicated vaginal birth | Not recommended | | | |
| Routine use of laxatives for the prevention of postpartum constipation | Not recommended | | | |
| Counseling and support for exclusive breastfeeding | Recommended | Yes | | Yes |
| Criteria to be assessed prior to discharge from the health facility after birth | Recommended | Yes | | Yes |
| Approaches to strengthen preparation for discharge from the health facility to home after birth (counselling) | Recommended | Yes | | Yes |
| Schedules for postnatal care contacts | Recommended | No | | |
| Length of stay in health facilities after birth | Recommended | No | | |
| Home visits for postnatal care contacts | Recommended | No | | |
| Midwifery continuity of care | Context-specific recommendation | No | | |
| Task sharing components of postnatal care delivery | Recommended | No | | |
| Recruitment and retention of staff in rural and remote areas | Recommended | No | | |
| Involvement of men in postnatal care and maternal and newborn health | Recommended with targeted | Yes | | No |

| Intervention | Recommended intervention | Clinical intervention | Level of care | HFA data available |
|---|---|---|---|---|
| | monitoring and evaluation | | | |
| Home-based records | Recommended | No | | |
| Digital targeted client communication | Context-specific recommendation | No | | |
| Digital birth notifications | Context-specific recommendation | No | | |
| **Newborn PNC** | | | | |
| Assessment of the newborn for danger signs | Recommended | Yes | | Yes |
| Cord care- Dry cord care or chlorhexidine / Application of chlorhexidine to the umbilical cord stump for the prevention of neonatal infection | Recommended (Chlorhexidine-Context-specific) | Yes | | Yes |
| Immunization for the prevention of infections | Recommended | Yes | | Yes |
| Timing of first bath to prevent hypothermia and its sequelae | Recommended | Yes | | No |
| Universal screening for abnormalities of the eye | Recommended | Yes | | No |
| Universal screening for hearing impairment | Recommended | Yes | | No |
| Universal screening for neonatal hyperbilirubinemia | Recommended | Yes | | No |
| Sleeping position for the prevention of sudden infant death syndrome | Recommended | Yes | | No |
| Whole-body massage | Recommended | Yes | | No |
| Early childhood development | Recommended | Yes | | No |
| Protecting, promoting, and supporting breastfeeding in facilities providing maternity and newborn services | Recommended | Yes | | No |
| Neonatal vitamin A supplementation | Context-specific | Yes | | No |
| Vitamin D supplementation for breastfed, term infants | Context-specific | Yes | | No |
| Use of emollients for the prevention of skin conditions | Not recommended | | | |

| Intervention | Recommended intervention | Clinical intervention | Level of care | HFA data available |
|---|---|---|---|---|
| **Small and sick newborn care** | | | | |
| Immediate newborn care (thorough drying, skin-to-skin contact of the newborn with the mother, delayed cord clamping, hygienic cord care) and Routine care (Vitamin K, eye care and vaccinations, weighing and clinical examinations) | | | Primary/Routine and essential newborn care | Yes |
| Early initiation and support for breastfeeding | | | Primary/Routine and essential newborn care | Yes |
| Prevention of mother-to-child transmission of HIV | | | Primary/Routine and essential newborn care | Yes |
| Neonatal resuscitation | | | Primary/Routine and essential newborn care | Yes |
| Pre-discharge advice on mother and baby care and follow up | | | Primary/Routine and essential newborn care | No |
| Thermal care | | | Secondary/Special newborn care | No |
| Kangaroo mother care, including follow-up | | | Secondary/Special newborn care | Yes |
| Comfort and pain management | | | Secondary/Special newborn care | Yes |
| Assisted feeding for optimal nutrition (cup feeding and nasogastric feeding) | | | Secondary/Special newborn care | No |
| Safe administration of oxygen | | | Secondary/Special newborn care | No |
| Prevention of apnea | | | Secondary/Special newborn care | No |
| Detection and management of neonatal infection (injectable antibiotics) | | | Secondary/Special newborn care | Yes |

| Intervention | Recommended intervention | Clinical intervention | Level of care | HFA data available |
| --- | --- | --- | --- | --- |
| Detection and management of hypoglycemia | | | Secondary/Special newborn care | Yes |
| Detection and management of jaundice (phototherapy) | | | Secondary/Special newborn care | No |
| Detection and management of anemia, including blood transfusion | | | Secondary/Special newborn care | No |
| Detection and management of neonatal encephalopathy | | | Secondary/Special newborn care | No |
| Seizure management | | | Secondary/Special newborn care | No |
| Safe administration of intravenous fluids | | | Secondary/Special newborn care | No |
| Detection and referral management of birth defects | | | Secondary/Special newborn care | No |
| Continuous positive airway pressure (CPAP) | | | Secondary/ Transition to intensive care | |
| Exchange transfusion | | | Secondary/ Transition to intensive care | |
| Detection and management of necrotizing enterocolitis (NEC) | | | Secondary/ Transition to intensive care | |
| Specialized follow up of high-risk infants (including preterm) | | | Secondary/ Transition to intensive care | |
| Advanced feeding support (e.g., parenteral nutrition) | | | Tertiary/Intensive newborn care | |
| Mechanical/assisted ventilation, including intubation | | | Tertiary/Intensive newborn care | |

| Intervention | Recommended intervention | Clinical intervention | Level of care | HFA data available |
|---|---|---|---|---|
| Screening and treatment for retinopathy of prematurity | | | Tertiary/Intensive newborn care | |
| Surfactant treatment | | | Tertiary/Intensive newborn care | |
| Investigation and management of birth defects | | | Tertiary/Intensive newborn care | |
| Pediatric surgery | | | Tertiary/Intensive newborn care | |
| Genetic services | | | Tertiary/Intensive newborn care | |

*Table S2: Guideline extraction*

| Intervention | Description of intervention | Equipment and supplies | Medicines and commodities | Diagnostics | Guidelines and staff training | Basic amenities | Routine service | Assessment | Intervention | Documentation and referral | Experience of care |
|---|---|---|---|---|---|---|---|---|---|---|---|
| **Maternal PNC** | | | | | | | | | | | |
| Physiological assessment of the woman | - Vaginal bleeding, uterine tonus, fundal height, temperature, and heart rate assessed during first 24 hour after birth<br>- Uterine void documented within 6 hours<br>- Blood pressure taken after birth, if normal second measure taken within six hours<br>- At each visit assess wellbeing of mother: micturition and urinary incontinence, bowel function, healing of any perineal wound, headache, fatigue, back pain, perineal pain and perineal hygiene, breast pain and uterine tenderness and lochia | (1) thermometer<br>(2) stethoscope<br>(3) blood pressure apparatus<br>(4) gloves<br>(5) soap and water for handwashing/ alcohol based handrub<br>(6) environmental disinfectant<br>(7) non-infectious waste (pedal bin receptable with lid and plastic liner) | | | (1) staff trained in maternal PNC<br>(2) available guidelines in maternal PNC | (1) bed for the mother<br>(2) room with auditory and visual privacy<br>(3) light source | | (1) assess for vaginal bleeding<br>(2) assess for uterine contractions<br>(3) assess fundal height<br>(4) measure temperature<br>(5) measure heart rate<br>(6) measure blood pressure taken after birth, if normal second measure taken within six hours<br>(7) assess urine void, micronutrition and urinary incontinence<br>(8) assess bowel function<br>(9) assess healing of any perineal wound<br>(10) assess headache<br>(11) assess fatigue<br>(12) assess back pain<br>(13) assess perineal pain and perineal hygiene | | | |

| Intervention | Description of intervention | Equipment and supplies | Medicines and commodities | Diagnostics | Guidelines and staff training | Basic amenities | Routine service | Assessment | Intervention | Documentation and referral | Experience of care |
|---|---|---|---|---|---|---|---|---|---|---|---|
| | | | | | | | | (14) assess breast pain (15) assess uterine tenderness and lochia (16) assess breastfeeding progress (17) Assess emotional well-being and how they are being supported (18) Ask about resolution of milk, transitory postpartum depression (19) Assess risk, signs and symptoms of domestic abuse (20) Ask about resumption of sexual intercourse and possible dyspareunia | | | |
| Prevention of postpartum depression and anxiety | All women would benefit from psychosocial interventions such as psychoeducation to develop coping strategies, manage stress and build supportive networks, where feasible and with availability of resources | | | | (1) staffed trained in delivering psychological support (2) available guidelines for the provision of psychological support | (1) room with auditory and visual privacy | | (1) assess for risk of postpartum depression (previous postpartum depression, previous mental illness, vulnerable population, traumatic childbirth, | (1) provide counselling and support to reduce risk of post-partum depression | | |

| Intervention | Description of intervention | Equipment and supplies | Medicines and commodities | Diagnostics | Guidelines and staff training | Basic amenities | Routine service | Assessment | Intervention | Documentation and referral | Experience of care |
|---|---|---|---|---|---|---|---|---|---|---|---|
| | | | | | | | | infant born preterm, stillbirth or neonatal death, infant admitted to intensive care and history of being a neglected child) | | | |
| Local cooling for perineal pain relief | Local cooling, such as with ice packs or cold pads, can be offered for the relief of acute pain from perineal trauma sustained during childbirth | (1) ice pack or crushed ice in a bag (gloves may also be used)<br>(2) perineal pad, gauze, cotton, or other skin barrier<br>(3) fresh, clean water and portable sitz bath or similar<br>(4) refrigeration | | | (1) staffed trained in maternal PNC<br>(2) available guidelines for maternal PNC | (1) electricity | | | (1) provision of cooling relief for pain | | |
| Oral analgesia for perineal pain relief | Oral paracetamol provided for relief of postpartum perineal pain | | (1) analgesic drugs (paracetamol) | | (1) staffed trained in administering paracetamol<br>(2) available guidelines for the provision of paracetamol | | | | (1) administer oral paracetamol | | |
| Pharmacological relief of pain due to uterine cramping/ involution | Oral non-steroidal anti-inflammatory drugs can be used for relief postpartum pain due to uterine cramping after birth | | (1) oral non-steroidal anti-inflammatory drugs (NSAIDs) | | (1) staffed trained in administering NSAIDS<br>(2) available guidelines | | | | (1) administer oral NSAIDs | | |

| Intervention | Description of intervention | Equipment and supplies | Medicines and commodities | Diagnostics | Guidelines and staff training | Basic amenities | Routine service | Assessment | Intervention | Documentation and referral | Experience of care |
|---|---|---|---|---|---|---|---|---|---|---|---|
| | | | | | for the provision of NSAIDS | | | | | | |
| Non-pharmacological interventions to treat postpartum breast engorgement | Counselling and support to practice responsive breastfeeding, good positioning, and attachment of the baby to the breast, expression of breastmilk, and the use of warm or cold compresses, based on a woman's preferences. | (1) cup, spoon or feeding bottle (2) gloves (3) soap and water for handwashing/alcohol based handrub (4) non-sharps waste (pedal bin receptable with lid and plastic liner) | | (1) warm/cold compress | (1) clear written breastfeeding policy (2) staff trained in breastfeeding support | (1) place for baby to lay while rooming in | | | (1) facilitate early and uninterrupted skin-to-skin contact between mothers and infants as soon as possible after birth (2) counselling and support for breastfeeding initiation and resolving common breastfeeding difficulties (3) counselling, coaching/support to express milk (4) counselling on responsive feeding (5) counselling about exclusive breastfeeding (6) counselling mothers on | | (1) respect the women's own thoughts, beliefs, and culture |

| Intervention | Description of intervention | Equipment and supplies | Medicines and commodities | Diagnostics | Guidelines and staff training | Basic amenities | Routine service | Assessment | Intervention | Documentation and referral | Experience of care |
|---|---|---|---|---|---|---|---|---|---|---|---|
| | | | | | | | | | avoidance of pacifiers (7) breastfeeding planning prior to discharge and linkage to continuing breastfeeding support (8) provide cold/warm compress | | |
| Non-pharmacological interventions to prevent postpartum mastitis | Counselling and support to practice responsive breastfeeding, good positioning, and attachment of the baby to the breast, expression of breastmilk, and the use of warm or cold compresses, based on a woman's preferences. | (1) cup, spoon or feeding bottle (2) gloves (3) soap and water for handwashing/alcohol based handrub (4) non-sharps waste (pedal bin receptable with lid and plastic liner) | | (1) warm/cold compress | (1) clear written breastfeeding policy (2) staff trained in breastfeeding support | (1) place for baby to lay while rooming in | | | (1) facilitate early and uninterrupted skin-to-skin contact between mothers and infants as soon as possible after birth (2) counselling and support for breastfeeding initiation and resolving common breastfeeding difficulties (3) counselling, coaching/support to express milk (4) counselling on | | (1) respect the women's own thoughts, beliefs, and culture |

| Intervention | Description of intervention | Equipment and supplies | Medicines and commodities | Diagnostics | Guidelines and staff training | Basic amenities | Routine service | Assessment | Intervention | Documentation and referral | Experience of care |
|---|---|---|---|---|---|---|---|---|---|---|---|
| | | | | | | | | | responsive feeding (5) counselling about exclusive breastfeeding (6) counselling mothers on avoidance of pacifiers (7) breastfeeding planning prior to discharge and linkage to continuing breastfeeding support (8) provide cold/warm compress | | |
| Prevention of postpartum constipation via dietary advice | Dietary advice and information on factors association with postpartum constipation | | | | (1) staff trained in dietary counselling (2) available guidelines for dietary counselling | | | | (1) provide dietary counselling | | |
| Screening for postpartum depression and anxiety | Use validated instrument to screen for postpartum depression. Should be accompanied by diagnostic and management services for women who screen positive. | (1) screening measures/tool/questionnaire | | | (1) staff trained in screening for postpartum depression (2) | | | (1) screen women for postpartum depression | | (1) refer/follow-up diagnostic and management for those | |

| Intervention | Description of intervention | Equipment and supplies | Medicines and commodities | Diagnostics | Guidelines and staff training | Basic amenities | Routine service | Assessment | Intervention | Documentation and referral | Experience of care |
|---|---|---|---|---|---|---|---|---|---|---|---|
| | | | | | available guidelines for screening of postpartum depression | | | | | who screen positive | |
| Physical activity and sedentary behavior | Provide clinical guidance to support women in participating in regular physical activity | | | | (1) staff trained in dietary counselling (2) available guidelines for dietary counselling | | | | (1) provide counselling and support to promote physical activity during postnatal care | | |
| Postpartum contraception | Provision of comprehensive contraceptive information and services during postnatal care including comprehensive contraceptive information, education, and counselling; and appropriate contraceptives | | (1) contraceptives (progestogen-only pills, progestogen-only injectable contraceptives, levonorgestrel, etonogestrel implants, copper-bearing intrauterine devices, levonorgestrel-releasing intrauterine devices, combined contraceptive patch, combined contraceptive | | (1) staff trained in the provision of contraceptives (2) available guidelines for the provision of contraceptives | | | (1) identification of appropriate contraceptive method | (1) provide contraceptives (2) provide adequate counselling about contraceptives | | |

| Intervention | Description of intervention | Equipment and supplies | Medicines and commodities | Diagnostics | Guidelines and staff training | Basic amenities | Routine service | Assessment | Intervention | Documentation and referral | Experience of care |
|---|---|---|---|---|---|---|---|---|---|---|---|
| | | | vaginal ring, progesterone-releasing vaginal ring, condoms, diaphragm, cervical cap, combined hormonal contraception ) | | | | | | | | |
| Postpartum oral iron and folate supplementation | Oral supplementation of iron supplementation, either with or without folic acid may be provided to postpartum women for 6-12 weeks post-delivery in settings where gestation anemia is of public health concern | (1) behavior change materials/counselling materials | (1) supplement containing iron dose and whether consumed daily or weekly should follow that used during pregnancy, or alternatively should start with that planned for menstruating women (daily oral iron and folic acid supplementation with 30 mg to 60 mg of elemental iron and 400 g (0.4 mg) of folic acid OR intermittent oral iron and folic acid supplementation with 120 mg of | | (1) staff trained in nutrition counselling (2) staff trained in provision and supplemental dosing of IFA (3) available guidelines on daily IFA supplementation during pregnancy | | | | (1) Provide/prescribe IFA supplement of: Daily IFA with 30-60mg elemental iron + 400ug folic acid (2) Provide, explain, or counsel on: instructions on correct dosing (3) Provide, explain, or counsel on: purpose and importance of IFA (4) Provide, explain, or counsel on: advise on potential side effects | (1) documentation of supplement provision | (1) The provision of effective clinical practices, relevant and timely information, psychosocial and emotional support by knowledgeable, supportive, and respective staff |

| Intervention | Description of intervention | Equipment and supplies | Medicines and commodities | Diagnostics | Guidelines and staff training | Basic amenities | Routine service | Assessment | Intervention | Documentation and referral | Experience of care |
|---|---|---|---|---|---|---|---|---|---|---|---|
| | | | elemental iron and 2800 g (2.8 mg)) | | | | | | | | |
| HIV catch-up testing | **In high burden areas:** HIV testing postpartum for women who are HIV negative or unknown status who missed early ANC contact testing or retesting in late pregnancy at a third trimester visit<br><br>**In low burden areas:** HIV testing postpartum for women who are HIV negative or unknown status who missed early ANC contact testing or retesting in late pregnancy at a third trimester visit. Could be conducted only for women in serodiscordant relationships or have had ongoing HIV risks in late pregnancy | IPC:<br>(1) gloves<br>(2) soap and water for handwashing/alcohol based handrub<br>(3) environmental disinfectant<br>(4) non-infectious waste (pedal bin receptable with lid and plastic liner)<br>(5) sharps container<br>(6) infectious waste container<br>For RDT:<br>(1) lancets<br>(2) alcohol swabs<br>(3) cotton wool<br>For ELISA:<br>(1) needle and syringe for blood draw | | (1) RDT kit or ELISA test (includes ELISA washer, ELISA reader, incubator, specific assay kit) | (1) guidelines on HIV counselling and testing<br>(2) staff trained on HIV counselling and testing | (1) room with auditory and visual privacy | | (1) conduct HIV testing | | (1) refer for follow-up/diagnostic and management services for those HIV positive | |
| Screening for tuberculosis disease | **In high burden areas**: systematic TB screening amongst the general population<br><br>**In low burden areas**: systematic TB screening amongst the general population may be conducted<br><br>Household contacts/close contacts of those with TB disease should be systematically screened | (1) TB register for contact tracing<br>(2) sputum collection container<br>(3) gloves<br>(4) soap and water for handwashing/alcohol based handrub<br>(5) environmental disinfectant<br>(6) infectious waste container<br>(7) non-infectious waste (pedal bin | | (1) (light or fluorescent microscope, slides, ZN stain) OR (fluorescent microscope, slides, and auramine-rhodamine stain) OR (observed system for sending sputum outside | (1) guidelines for diagnosis of TB<br>(2) staff trained in diagnosis of TB | | | (1) conduct TB testing | | (1) refer for follow-up/diagnostic and management services for those with TB | |

| Intervention | Description of intervention | Equipment and supplies | Medicines and commodities | Diagnostics | Guidelines and staff training | Basic amenities | Routine service | Assessment | Intervention | Documentation and referral | Experience of care |
|---|---|---|---|---|---|---|---|---|---|---|---|
| | | receptable with lid and plastic liner) | | facility for TB diagnosis and receiving results) | | | | | | | |
| Preventive antihelminthic treatment | Provision of annual or biannual preventive chemotherapy (albendazole or mebendazole) in areas where baseline prevalence of soil-transmitted helminth infection if 20+ or more | | (1) single dose albendazole (400mg) or mebendazole (500mg) | | (1) staffed trained in administering chemotherapy (2) available guidelines for chemotherapy | | | | (1) provide deworming | | |
| Preventive schistosomiasis treatment | Annual preventive chemotherapy with praziquantel in endemic communities with Schistosoma prevalence >10% | | (1) praziquantel | | (1) staffed trained in administering chemotherapy (2) available guidelines for chemotherapy | | | | (1) provide praziquantel | | |
| Oral pre-exposure prophylaxis for HIV prevention | Oral PrEP containing tenofovir disoproxil fumarate should be started or continued postpartum for women at substantial risk of HIV infection | | (1) PrEP containing tenofovir disoproxil fumarate | | (1) staff trained in administering PrEP (2) available guidelines for administering PrEP | | | (1) identification of women at substantial risk of HIV infection | (1) provide PrEP to women (2) counselling to support PrEP use | | |
| Exclusive breastfeeding | Counselling and support provided to women (WHO guidelines); rooming in 24 hours a day. Additional | (1) cup, spoon or feeding bottle (2) gloves (3) soap and water | | | (1) clear written breastfeeding policy | (1) place for baby to lay while rooming in | | | (1) facilitate early and uninterrupted skin-to- | | (1) respect the women's own |

| Intervention | Description of intervention | Equipment and supplies | Medicines and commodities | Diagnostics | Guidelines and staff training | Basic amenities | Routine service | Assessment | Intervention | Documentation and referral | Experience of care |
|---|---|---|---|---|---|---|---|---|---|---|---|
| | support provided for pre-term babies and low birth weight babies.<br><br>Includes: non-pharmacological intervention to treat postpartum breast engorgement, and non-pharmacological interventions to prevent postpartum mastitis | for handwashing/alcohol based handrub (4) non-sharps waste (pedal bin receptable with lid and plastic liner) | | | (2) staff trained in breastfeeding support | | | | skin contact between mothers and infants as soon as possible after birth (2) counselling and support for breastfeeding initiation and resolving common breastfeeding difficulties (3) counselling, coaching/support to express milk (4) counselling on responsive feeding (5) counselling about exclusive breastfeeding (6) counselling mothers on avoidance of pacifiers (7) oral stimulation for pre-term babies who are not able | | thoughts, beliefs, and culture |

| Intervention | Description of intervention | Equipment and supplies | Medicines and commodities | Diagnostics | Guidelines and staff training | Basic amenities | Routine service | Assessment | Intervention | Documentation and referral | Experience of care |
|---|---|---|---|---|---|---|---|---|---|---|---|
| | | | | | | | | | to breastfeed (8) breastfeeding planning prior to discharge and linkage to continuing breastfeeding support | | |
| Criteria to be assessed prior to discharge from the health facility after birth | Prior to discharging from the health facility, health workers should assess:<br>- the woman's and baby's physical well-being and the woman's emotional well-being;<br>- the skills and confidence of the woman to care for herself and the skills and confidence of the parents and caregivers to care for the newborn; and<br>- the home environment and other factors that may influence the ability to provide care for the woman and the newborn in the home, and care-seeking behavior | (1) behavior change materials/counselling materials | | | (1) staff trained in maternal PNC<br>(2) available guidelines in maternal PNC | (1) room with auditory and visual privacy | | (1) assessment of the mother's wellbeing<br>(2) assessment of the mother's skills and confident to care for newborn<br>(3) assessment of the home environment and other factors that will influence newborn care and care-seeking behaviors | | | |
| Approaches to strengthen preparation for discharge from the health facility to home after birth (counselling) | Provide information, education, and counselling to prepare women, parents, and caregivers for discharge from health facility to improve maternal and newborn health outcomes | (1) education materials<br>(2) job aids for providers | | | (1) staff trained in maternal PNC<br>(2) available guidelines in maternal PNC | (1) room with auditory and visual privacy | | | (1) counselling about physical process of recovery after birth<br>(2) counselling about signs and symptoms of post-partum | | |

| Intervention | Description of intervention | Equipment and supplies | Medicines and commodities | Diagnostics | Guidelines and staff training | Basic amenities | Routine service | Assessment | Intervention | Documentation and referral | Experience of care |
|---|---|---|---|---|---|---|---|---|---|---|---|
| | | | | | | | | | hemorrhage (3) counselling about signs and symptoms of pre-eclampsia/eclampsia (4) counselling about signs and symptoms of infection (5) counselling about signs and symptoms of thromboembolism (6) counselling about nutrition (7) counselling about hygiene, especially handwashing (8) counselling about birth spacing and family planning, discuss contraceptive and provide contraceptives if | | |

| Intervention | Description of intervention | Equipment and supplies | Medicines and commodities | Diagnostics | Guidelines and staff training | Basic amenities | Routine service | Assessment | Intervention | Documentation and referral | Experience of care |
|---|---|---|---|---|---|---|---|---|---|---|---|
| | | | | | | | | | requested<br>(9) counselling about safer sex including use of condoms<br>(10) counselling about sleep under ITN<br>(11) counselling about gentle exercise | | |
| Involvement of men in postnatal care and maternal and newborn health | Interventions to promote the involvement of men during pregnancy, childbirth and after birth are recommended to facilitate and support improved self-care of women, home care practices for women and newborns, and use of skilled care for women and newborns during pregnancy, childbirth and the postnatal period, and to increase the timely use of facility care for obstetric and newborn complications.<br><br>Recommended interventions include:<br>- couples education – interventions that included educational activities with couples, conducted in the home or in a facility, with either an individual couple or in groups;<br>- men's education – educational activities directed towards men, | (1) education materials<br>(2) job aids for providers | | | (1) staff trained in provision of counselling and education to men involved in PNC<br>(2) available guidelines for the involvement of men in PNC | | | | (1) Provide education/counselling to males involved in PNC | | (1) promote the involvement of men in a way that respects, promotes, and facilitates women's choices and their autonomy in decision-making, and that supports women in taking care of themselves and their newborns |

| Intervention | Description of intervention | Equipment and supplies | Medicines and commodities | Diagnostics | Guidelines and staff training | Basic amenities | Routine service | Assessment | Intervention | Documentation and referral | Experience of care |
|---|---|---|---|---|---|---|---|---|---|---|---|
| | conducted in groups or individually, in the health facility or the community, or through text-messaging;<br>- multicomponent interventions that included either men only or couples education activities as well as community-mobilization, mass media efforts, home visits, etc.;<br>- having a companion during labor and birth, including having the father cut the umbilical cord after birth | | | | | | | | | | |
| **Newborn PNC** | | | | | | | | | | | |
| Assessment of the newborn for danger signs | Assessed and referred for further evaluations if:<br>- stopped feeding well<br>- history of convulsions<br>-fast breathing<br>-severe chest in-drawing<br>-no spontaneous movement<br>-fever<br>-lower body temperature<br>-any jaundice in the first 24 hours | (1) thermometer<br>(2) stethoscope<br>(3) soap and water for handwashing/alcohol based handrub<br>(4) gloves<br>(5) non-sharps waste (pedal bin receptable with lid and plastic liner) | | | (1) staff trained in newborn assessment<br>(2) available guidelines in newborn assessment | (1) room with auditory and visual privacy | | (1) ask if the baby is feeding well and/or observe breastfeeding<br>(2) ask if the baby has had convulsions<br>(3) assess breathing (listen for grunting, count breaths for 1 minute)<br>(4) look at chest for in-drawing<br>(5) look at the babies movements/try to wake the baby if sleeping<br>(6) assess babies temperature<br>(7) look at color of skin | | | |

| Intervention | Description of intervention | Equipment and supplies | Medicines and commodities | Diagnostics | Guidelines and staff training | Basic amenities | Routine service | Assessment | Intervention | Documentation and referral | Experience of care |
|---|---|---|---|---|---|---|---|---|---|---|---|
| | | | | | | | | for signs of jaundice in first 24 hours<br>(8) look at color of palms and soles for jaundice after first 24 hours | | | |
| Cord care- Dry cord care or Chlorhexidine / Application of chlorhexidine to the umbilical cord stump for the prevention of neonatal infection | **Dry cord care**: clean dry cord care recommended for newborns born in health facility;<br>If stump is soiled, wash it with clean water and soap. Dry it thoroughly with clean cloth.<br>**Chlorhexidine**: daily chlorhexidine application to the stump during the first week of life for babies born at home | (1) clean cloth<br>(2) soap and water<br>(3) environmental disinfectant<br>(4) non-sharps waste (pedal bin receptable with lid and plastic liner) | (1) 7.1% chlorhexidine digluconate aqueous solution or gel, delivering 4% chlorhexidine | | (1) staff trained in provision of cord care<br>(2) available guidelines on cord care practices | | | | (1) counselling or instructions on cord care<br>(2) application of chlorhexidine | | |
| Immunization for the prevention of infections | Give BCG, OPV-0, Hepatitis B vaccine birth dose, within 24 hours after birth, preferably before discharge. | (1) needle + syringe<br>(2) child health card/vaccine card<br>(3) immunization tally sheets<br>(4) cold box/vaccine carrier with ice packs<br>(5) refrigerator<br>(6) temperature monitoring device in refrigerator<br>(7) sterile water<br>(8) intradermal needle (more common) OR multiple puncture device (BCG)<br>(9) sharps container<br>(10) gloves | (1) BCG vaccine<br>(2) Hepatitis B vaccine<br>(3) OPV-0 vaccine | | (1) staff trained in child immunization<br>(2) available guidelines for child immunization | | | | (1) delivery of BCG vaccine<br>(2) delivery of Hep B vaccine<br>(3) delivery of polio vaccine<br>(4) advise to return for next dose | (1) documentation of BCG vaccine<br>(2) documentation of Hep B vaccine<br>(3) documentation of polio vaccine | |

| Intervention | Description of intervention | Equipment and supplies | Medicines and commodities | Diagnostics | Guidelines and staff training | Basic amenities | Routine service | Assessment | Intervention | Documentation and referral | Experience of care |
|---|---|---|---|---|---|---|---|---|---|---|---|
| | | (11) pedal bin receptable with lid and plastic liner)<br>(12) power<br>(13) soap and water for handwashing/alcohol based handrub | | | | | | | | | |
| Timing of first bath to prevent hypothermia and its sequelae | Immediately drying and wrapping (appropriate clothes and hat/caps) or skin to skin, no bathing for the first 24 hours (if not possible because of cultural reasons should be delayed for 6 hours) | (1) 2 or more absorbent towels<br>(2) clothes to wrap baby<br>(3) hat/caps<br>(4) sheet or blanket to cover mother and baby<br>(5) bedding<br>(6) warm water<br>(7) dry towel<br>(8) education materials for staff<br>(9) education materials for parents | | | (1) staff trained in thermal care<br>(2) staff trained in early breastfeeding<br>(3) available guidelines for thermal care<br>(4) available guidelines for early breastfeeding | | | | (1) immediately dry baby after birth<br>(2) wrap the baby and put on hat/cap<br>(3) promote skin-to-skin<br>(4) early initiation of breastfeeding<br>(5) delayed bathing | | |
| Universal screening for abnormalities of the eye | External examination of the eye and red reflect test using standard equipment should be conducted prior to discharge. | (1) information materials for parents<br>(2) batteries<br>(3) medical flashlight/penlight<br>(4) ophthalmoscope set | | | (1) staff trained in universal screening for abnormalities of the eye<br>(2) available guidelines for universal screening for | (1) darkened room/space | | (1) conduct external examination of the eye and red reflect test | | (1) refer for follow-up/diagnostic and management services for children identified with abnormality | |

| Intervention | Description of intervention | Equipment and supplies | Medicines and commodities | Diagnostics | Guidelines and staff training | Basic amenities | Routine service | Assessment | Intervention | Documentation and referral | Experience of care |
|---|---|---|---|---|---|---|---|---|---|---|---|
| | | | | | abnormalities of the eye | | | | | | |
| Universal screening for hearing impairment | Universal newborn hearing screening with otoacoustic emissions or automated auditory brainstem response for early identification of permanent bilateral hearing loss. | (1) information materials for parents<br>(2) OAE device and software including small outer-ear probe with earphone, microphone, and ear tip OR<br>(3) AABR device and software with ear coupler, and disposable earphones and sensors/electrodes | | | (1) staff trained in universal newborn hearing screening<br>(2) available guidelines for universal newborn hearing screening | (1) quiet calm space<br>(2) electricity | | (1) conduct hearing screening with otoacoustic emission or automated auditory brainstem response | | (1) refer for follow-up/diagnostic and management services for children identified with hearing loss | |
| Universal screening for neonatal hyperbilirubinemia | Universal screening for neonatal hyperbilirubinemia by transcutaneous bilirubinometer | (1) alcohol wipes<br>(2) reusable probe tips<br>(3) printed nomogram OR (4) computer for patient monitoring<br>(5) non-sharps waste | | (1) TcB (transcutaneous bilirubinometer OR icterometer) | (1) staff trained in screening for neonatal hyperbilirubinemia by transcutaneous bilirubinometer<br>(2) available guidelines for screening for neonatal hyperbilirubinemia by transcutaneous | (1) electricity | | (1) conduct screening for neonatal hyperbilirubinemia by transcutaneous | | (1) refer for follow-up/diagnostic and management services for children identified with neonatal hyperbilirubinemia | |
| Sleeping position for the prevention of | Counselling on putting baby to sleep in the supine | (1) information materials for | | | (1) staff trained to | | | | (1) counselling | | |

| Intervention | Description of intervention | Equipment and supplies | Medicines and commodities | Diagnostics | Guidelines and staff training | Basic amenities | Routine service | Assessment | Intervention | Documentation and referral | Experience of care |
|---|---|---|---|---|---|---|---|---|---|---|---|
| sudden infant death syndrome | position during the first year of life | parents<br>(2) place for a baby to lie | | | support supine sleeping<br>(2) available guidelines about supine sleeping | | | | on sleep position | | |
| Whole-body massage | Provision of gentle whole-body massage for term healthy newborn | (1) instructional brochures, videos or similar<br>(2) soap and water for handwashing/alcohol based handrub | (1) massage oil (optional) | | (1) staff trained in the infant massage<br>(2) available guidelines about infant massage | | | | (1) conduct whole body massage | | |
| Early childhood development | Responsive feeding<br>Engagement with early learning activities | | | | (1) staff trained in responsive feeding OR staff trained in early learning<br>(2) available guidelines on encouraging play and communication | | | | (1) counselling to encourage communication and play<br>(2) counselling to improve responsive feeding | | |
| Protecting, promoting, and supporting breastfeeding in facilities providing maternity and newborn services | Facilities providing maternity and newborn services should have a clearly written breastfeeding policy that is routinely communicated to staff and parents<br>Health-facility staff who | | | | (1) clear written breastfeeding policy<br>(2) staff trained in breastfeed | | | | | | |

| Intervention | Description of intervention | Equipment and supplies | Medicines and commodities | Diagnostics | Guidelines and staff training | Basic amenities | Routine service | Assessment | Intervention | Documentation and referral | Experience of care |
|---|---|---|---|---|---|---|---|---|---|---|---|
| | provide infant feeding services, including breastfeeding support, should have sufficient knowledge, competence, and skills to support women to breastfeed | | | | ing support | | | | | | |
| Neonatal vitamin A supplementation | Provide newborn with an oral dose of 50 000 IU for vitamin A within the first 3 days after birth in settings with high infant mortality rates and high prevalence of maternal vitamin A deficiency | (1) scissors<br>(2) non-sharps waste<br>(3) refrigerators (in some settings) | (1) vitamin A (50,000IU/ml drops) | | (1) staff trained in the provision of neonatal vitamin A<br>(2) available guidelines about the provision of neonatal vitamin A | | | | (1) provision of vitamin A | (1) documented provision of vitamin A | |
| Vitamin D supplementation for breastfed, term infants | Vitamin D supplementation within the first week | (1) refrigerators (in some settings) | (1) vitamin D3 drops (10 000 IU/mL) | | (1) staff trained in the provision of vitamin D<br>(2) available guidelines about the provision of vitamin D | | | | (1) provide vitamin D | | |
| **Small and sick newborn care** | | | | | | | | | | | |
| Immediate newborn care and Routine care | Immediate newborn care includes thorough drying, skin-to-skin contact of the newborn with the mother, delayed cord clamping, hygienic cord care. Routine care includes Vitamin K, eye | (1) clean, dry blankets/towels (for drying baby)<br>(2) linen (to wrap baby)<br>(3) sterile scissors and/or sterile | (1) vitamin k<br>(2) antibiotic eye ointment for newborns (tetracycline or other)<br>(3) | | (1) staff trained in clean cord cutting and appropriate cord care | (1) electricity supply | (1) routine, complete (head-to-toe) examination of newborn | | | | |

| Intervention | Description of intervention | Equipment and supplies | Medicines and commodities | Diagnostics | Guidelines and staff training | Basic amenities | Routine service | Assessment | Intervention | Documentation and referral | Experience of care |
|---|---|---|---|---|---|---|---|---|---|---|---|
| | care and vaccinations, weighing and clinical examinations | blade to cut cord<br>(4) umbilical cord clamp (sterile ligatures or clamp of Barr) or cord ties/sterile thread<br>(5) infant scale<br>(6) radiant heater, warmth source<br>(7) newborn caps<br>(8) newborn socks<br>(9) single use needles and syringes<br>(10) alcohol swabs<br>(11) cotton wool<br>(12) thermometer for low body temperature<br>(13) stethoscope<br>(14) refrigerator for vaccines with<br>(15) temperature monitoring device in refrigerator or<br>(16) cold box/vaccine carrier with ice packs<br>(17) non-sharps waste container (pedal bin receptable with lid and plastic liner)<br>(18) disinfectant solutions (e.g. chlorine bleach)<br>(19) gloves (disposable)<br>(20) sharps container<br>(21) soap and water for handwashing or | chlorhexidine solution (country specific based on national policy)<br>(4) BCG vaccine<br>(5) oral poliomyelitis vaccine<br>(6) hep B vaccine | | (2) staff trained in thermal care<br>(3) essential newborn care guidelines<br>(4) national immunization schedule | | before discharge | | | | |

| Intervention | Description of intervention | Equipment and supplies | Medicines and commodities | Diagnostics | Guidelines and staff training | Basic amenities | Routine service | Assessment | Intervention | Documentation and referral | Experience of care |
|---|---|---|---|---|---|---|---|---|---|---|---|
| | | alcohol-based hand rub | | | | | | | | | |
| Early initiation and support for breastfeeding | Early initiation and support for breastfeeding | (1) gloves (disposable)<br>(2) soap and water for handwashing or alcohol-based hand rub | | | (1) staff trained in early and exclusive breastfeeding<br>(2) educational information on breastfeeding for mothers (e.g., written and pictorial information, support classes or groups, posters)<br>(3) clear written breastfeeding policy | | (1) initiation of breastfeeding within the first hour | | | | |
| Prevention of mother-to-child transmission of HIV | Early infant diagnosis and rapid enrollment of infants living with HIV in care and treatment programs | (1) gloves (disposable)<br>(2) soap and water for handwashing or alcohol based handrub<br>(3) non-sharps waste container (pedal bin receptable with lid and plastic liner)<br>(4) sharps container<br>(5) needle and syringe for blood | (1) antiretrovirals for babies (nevirapine syrup or zidovudine syrup)<br>(2) antiretrovirals for mothers (option a: azt, nvp, and 3tc option b: azt + 3tc + lpv or azt + 3tc + abc | (1) HIV diagnostic capacity / HIV nucleic-acid testing | (1) PMTCT guidelines<br>(2) infant and young child feeding counseling guidelines<br>(3) staff trained ARV prophylactic treatment for PMTCT | (1) PMTCT room is private room with auditory and visual privacy | | | | | |

| Intervention | Description of intervention | Equipment and supplies | Medicines and commodities | Diagnostics | Guidelines and staff training | Basic amenities | Routine service | Assessment | Intervention | Documentation and referral | Experience of care |
|---|---|---|---|---|---|---|---|---|---|---|---|
| | | draw<br>for heel prick:<br>(6) filter paper for DBS<br>(7) lancets<br>(8) alcohol swabs<br>(9) cotton wool | or azt + 3tc + efv or tdf + 3tc (or ftc) + efv)<br>(3) preventative therapy (cotrimoxazole) | | (4) staff trained newborn nutrition counseling of mother with HIV<br>(5) staff trained infant and young child feeding | | | | | | |
| Neonatal resuscitation | Emergency care is given to help a baby who is not breathing or who is making a lot of effort with their breathing | (1) airway suction apparatus (suction apparatus with catheter or suction bulb for mucus extraction)<br>(2) infant resuscitation bag/mask<br>(3) stethoscope<br>(4) nasal prongs 1mm and 2mm (if nasal prongs not available use nasal catheter (8-f and 6-f sizes)<br>(5) neonatal sized pulse oximetry probes/sensors for oxygen saturations<br>(6) consistent oxygen source/supply (e.g. oxygen concentrators)<br>(7) oxygen humidifiers<br>(8) oxygen low flow device<br>(9) oxygen flow | | | (1) staff trained in neonatal resuscitation using bag and mask<br>(2) BEmONC guidelines<br>(3) wall charts/action sequences for neonatal resuscitation (e.g., HBB flowchart) | (1) electricity supply | (1) facility past three months provided neonatal resuscitation | | | | |

| Intervention | Description of intervention | Equipment and supplies | Medicines and commodities | Diagnostics | Guidelines and staff training | Basic amenities | Routine service | Assessment | Intervention | Documentation and referral | Experience of care |
|---|---|---|---|---|---|---|---|---|---|---|---|
| | | splitter for newborn<br>(10) oxygen tubing<br>(11) apnea monitor<br>(12) oxygen blenders<br>(13) pulse oximeter<br>(14) soap and water for handwashing or alcohol-based hand rub<br>(15) gloves (disposable) | | | | | | | | | |
| Pre-discharge advice on mother and baby care and follow up | Small and sick newborns may require additional follow-up to assess recovery, feeding and weight gain and to monitor health and neurodevelopmental progress. Some newborns may require follow-up appointments with medical specialists (ophthalmology, neurology, cardiology, surgery) and allied health professionals (nutrition, speech therapy, physiotherapy, occupational therapy, audiology). Parents and carers should be educated and trained to build their confidence in caring for their newborns at home. | (1) gloves (disposable)<br>(2) soap and water for handwashing or alcohol-based hand rub | | | (1) the health facility has written, up-to-date guidelines, protocols and standard operating procedures for discharge of newborns, including a pre-discharge baby check, a comprehensive discharge management plan and arranging follow-up | (1) room with auditory and visual privacy | | | | | |

| Intervention | Description of intervention | Equipment and supplies | Medicines and commodities | Diagnostics | Guidelines and staff training | Basic amenities | Routine service | Assessment | Intervention | Documentation and referral | Experience of care |
|---|---|---|---|---|---|---|---|---|---|---|---|
| | | | | | as required.<br>(2) health care staff in the health facility who care for small and sick newborns receive in-service training and regular refresher sessions in discharge planning and follow-up<br>(3) behavior change materials/ counselling materials for mother and baby care | | | | | | |
| Thermal care | All small babies need attention to basic thermal care to prevent them from becoming cold. Assist mothers to provide skin-to-skin care for<br>small babies in the first 24 hours after birth.<br>• Dry the baby thoroughly at birth, cover the head, and place the baby skin-to-skin.<br>• Put on a diaper and dry head covering. | (1) 2 or more absorbent towels<br>(2) clean, dry blankets/towels (for drying baby)<br>(3) linen (to wrap baby)<br>(4) newborn socks<br>(5) newborn caps<br>(6) diapers<br>(7) gloves (disposable)<br>(8) soap and water | | | (1) counseling material on thermal care<br>(2) ENC/BEmONC or thermal care guidelines<br>(3) staff trained in | | (1) drying and wrapping newborns to keep them warm | | | | |

| Intervention | Description of intervention | Equipment and supplies | Medicines and commodities | Diagnostics | Guidelines and staff training | Basic amenities | Routine service | Assessment | Intervention | Documentation and referral | Experience of care |
|---|---|---|---|---|---|---|---|---|---|---|---|
| | | for handwashing or alcohol-based hand rub | | | ENC/BEmONC or thermal care | | | | | | |
| Kangaroo mother care | Care of preterm infants carried skin-to-skin with the mother. Its key features include early continuous and prolonged skin-to-skin contact between the mother and the baby, and exclusive breastfeeding (ideally) or feeding with breastmilk. | (1) clean, dry blankets/towels (for drying baby)<br>(2) linen (to wrap baby)<br>(3) newborn socks<br>(4) newborn caps<br>(5) support binder (pouch, shirt, band, fabric)<br>(6) diapers<br>(7) gloves (disposable)<br>(8) soap and water for handwashing or alcohol-based hand rub<br>(9) infant scale | | | (1) counseling material on KMC<br>(2) staff trained in KMC for low-birth-weight babies<br>(3) guidelines on KMC | (1) separate room or space for KMC<br>(2) private washing areas and toilet for mothers<br>(3) food provision for mothers/area for preparation of food<br>(4) sufficient space for mothers to store personal items, comfortable chairs, and privacy | (1) facility practices KMC | | | | |
| Comfort and pain management | Pain affects brain development, with potential long-term effects. Health care providers should recognize pain cues and know how to prevent and minimize pain in newborns. Appropriate tools are recommended for assessing pain and making decisions on pain management and comfort. Comfort is improved when newborns are encouraged | (1) blankets for swaddling newborn<br>(2) medication delivery mechanism (infusion kit + any IV fluid or single use syringes)<br>(3) gloves (disposable)<br>(4) soap and water for handwashing or alcohol-based | (1) oral sucrose<br>(2) paracetamol<br>(3) morphine (IV) | | (1) the health facility has written, up-to-date guidelines, protocols and standard operating procedures and tools for the assessmen | | | | | | |

| Intervention | Description of intervention | Equipment and supplies | Medicines and commodities | Diagnostics | Guidelines and staff training | Basic amenities | Routine service | Assessment | Intervention | Documentation and referral | Experience of care |
|---|---|---|---|---|---|---|---|---|---|---|---|
| | to breastfeed or placed skin-to-skin with a family member. Oral sucrose may be used during potentially painful procedures, such as blood sampling heel pricks. For sicker newborns, analgesics with appropriate monitoring may be necessary. | hand rub<br>(5) non-sharps waste container (pedal bin receptable with lid and plastic liner)<br>(6) sharps container<br>(7) disinfectant solutions (e.g. chlorine bleach) | | | t, recognition, prevention, and management of pain in small and sick newborns.<br>(2) the health staff receive training and regular refresher courses in assessing, preventing, and controlling pain in small and sick newborns. | | | | | | |
| Assisted feeding for optimal nutrition | Many preterm newborns have delayed or impaired sucking and require help, which may include expressing human milk, cup, or nasogastric tube feeding. If expressed breast milk or other feeds are required for preterm infants, feeding methods such as cups or spoons are preferable to feeding bottles and teats. | (1) feeding cups and spoons<br>(2) stethoscope<br>(3) adhesive tape/strapping for NG tubes<br>(4) bottles, teats, dummies (as appropriate for feeding guidelines)<br>(5) breast pumps (battery powered)<br>(6) collection containers (for expressed breastmilk) | (1) breastmilk substitute (only for babies with mothers unable to express milk) (i.e., human donor milk, infant formula)<br>(2) breast milk fortifier | | (1) educational information on breastfeeding for mothers (e.g. written and pictorial information, posters)<br>(2) the health facility has | (1) area or room for breastmilk expression<br>(2) milk room/area for preparing milk feeds and storage of expressed breastmilk<br>(3) electricity supply | | | | | |

| Intervention | Description of intervention | Equipment and supplies | Medicines and commodities | Diagnostics | Guidelines and staff training | Basic amenities | Routine service | Assessment | Intervention | Documentation and referral | Experience of care |
|---|---|---|---|---|---|---|---|---|---|---|---|
| | | (7) nasogastric feeding tubes 3.5-10 with caps<br>(8) refrigerator and freezer (for milk storage only)<br>(9) sterile feeding syringes (2.5ml, 5ml, 10ml)<br>(10) utensils and containers for preparing milk feeds especially graduated measuring jug/cup<br>(11) gloves (disposable)<br>(12) soap and water for handwashing or alcohol based hand rub | | | written, up-to-date guidelines, protocols and standard operating procedures for exclusive breastfeeding and optimal feeding of small and sick newborns, including newborns of HIV-infected mothers, that include target volumes, feeding advancement and criteria for initiating exclusive enteral nutrition, consistent with who guidelines.<br>(3) health care staff in the health facility who care for | | | | | | |

| Intervention | Description of intervention | Equipment and supplies | Medicines and commodities | Diagnostics | Guidelines and staff training | Basic amenities | Routine service | Assessment | Intervention | Documentation and referral | Experience of care |
|---|---|---|---|---|---|---|---|---|---|---|---|
| | | | | | newborns receive in-service training and regular refresher sessions in counselling on breastfeeding and optimal feeding of small and sick newborns, including newborns of HIV-infected mothers | | | | | | |
| Safe administration of oxygen | Small and sick newborns requiring supplemental oxygen therapy receive it safely through appropriate neonatal equipment, including neonatal nasal prongs, low-flow meters, air–oxygen blenders, humidifiers, and pulse oximeters. | (1) nasal prongs 1mm and 2mm (if nasal prongs not available use nasal catheter (8-f and 6-f sizes)<br>(2) neonatal sized pulse oximetry probes/sensors for oxygen saturations<br>(3) consistent oxygen source/supply (e.g. oxygen concentrators)<br>(4) oxygen humidifiers<br>(5) oxygen low flow device<br>(6) oxygen flow splitter for | | | (1) the health facility has written, up-to-date oxygen therapy guidelines, protocols and standard operating procedures for assessment of hypoxia and hyperoxia with a neonatal pulse | (1) electricity supply | | | | | |

| Intervention | Description of intervention | Equipment and supplies | Medicines and commodities | Diagnostics | Guidelines and staff training | Basic amenities | Routine service | Assessment | Intervention | Documentation and referral | Experience of care |
|---|---|---|---|---|---|---|---|---|---|---|---|
| | | newborn<br>(7) oxygen tubing<br>(8) apnea monitor<br>(9) oxygen blenders<br>(10) pulse oximeter<br>(11) soap and water for handwashing or alcohol-based hand rub<br>(12) gloves (disposable) | | | oximeter, determining the need for oxygen therapy and safe use of supplemental oxygen in small, sick and preterm newborns according to who guidelines.<br>(2) the health facility clinical staff who care for newborns receive training and regular refresher sessions in assessing and managing newborns with respiratory conditions | | | | | | |
| Prevention of apnea | Recurrent apnea (a pause in breathing of greater than 20 seconds) is common in preterm infants, particularly those at very early gestational ages. Such episodes of ineffective breathing can lead to | (1) medication delivery mechanism (infusion kit + any IV fluid or single use syringes)<br>(2) gloves (disposable) | (1) methylxanthines (such as caffeine, theophylline, and aminophylline; caffeine is | | (1) the health facility has written, up-to-date guidelines, protocols, and | | | | | | |

| Intervention | Description of intervention | Equipment and supplies | Medicines and commodities | Diagnostics | Guidelines and staff training | Basic amenities | Routine service | Assessment | Intervention | Documentation and referral | Experience of care |
|---|---|---|---|---|---|---|---|---|---|---|---|
| | hypoxia and bradycardia that may be severe enough to require positive pressure ventilation. Methylxanthines (such as caffeine, theophylline, and aminophylline) have been used to stimulate breathing and reduce apnea and its consequences. | (3) soap and water for handwashing or alcohol-based hand rub (4) non-sharps waste container (pedal bin receptable with lid and plastic liner) (5) sharps container (6) disinfectant solutions (e.g., chlorine bleach) | the preferred drug); IV delivery | | standard operating procedures for assessing, managing, and preventing apnea in newborns. (2) the health facility clinical staff who care for newborns receive training and regular refresher sessions in assessing and managing newborns with respiratory conditions | | | | | | |
| Detection and management of neonatal infection (injectable antibiotics) | All small and sick newborns are assessed for possible serious bacterial infections and receive prompt antibiotics and supportive care consistent with WHO guidance. | (1) medication delivery mechanism (infusion kit + any IV fluid or single use syringes) (2) infant scale (3) pulse oximeter (4) consistent oxygen source/supply (e.g. oxygen concentrators) (5) gloves | (1) first line antibiotic treatment (ampicillin (or penicillin) and gentamicin) (2) alternative treatment if at risk of staphylococcus infection (cloxacillin and gentamicin | (1) blood culture (2) c-reactive protein (CRP) (3) full blood count (FBC) (4) lumbar puncture (LP) (5) chest x-ray (CXR) | (1) staff trained in newborn infection management (including injectable antibiotics) (2) the health facility has written, up-to-date | (1) separate area/clean space for preparing IV drugs (can be the same as area for iv fluids) (2) electricity supply | | | | | |

| Intervention | Description of intervention | Equipment and supplies | Medicines and commodities | Diagnostics | Guidelines and staff training | Basic amenities | Routine service | Assessment | Intervention | Documentation and referral | Experience of care |
|---|---|---|---|---|---|---|---|---|---|---|---|
| | | (disposable)<br>(6) soap and water for handwashing or alcohol based hand rub<br>(7) non-sharps waste container (pedal bin receptable with lid and plastic liner)<br>(8) sharps container<br>(9) disinfectant solutions (e.g. chlorine bleach) | instead of penicillin and gentamicin)<br>(3) home-based care – amoxicillin (oral suspension) and intramuscular gentamicin | | guidelines, protocols and standard operating procedures consistent with who guidelines for prevention, early diagnosis and management of neonatal infection in the maternity unit and all areas where newborns are assessed and managed.<br>(3) the health facility has written, up-to-date guidelines, protocols and standard operating procedures on safe and rational use of | | | | | | |

| Intervention | Description of intervention | Equipment and supplies | Medicines and commodities | Diagnostics | Guidelines and staff training | Basic amenities | Routine service | Assessment | Intervention | Documentation and referral | Experience of care |
|---|---|---|---|---|---|---|---|---|---|---|---|
| | | | | | antibiotics and other medications for small and sick newborns, based on their weight or age. | | | | | | |
| Detection and management of hypoglycemia | Small and sick newborns are at risk of low blood glucose. These babies often have limited glycogen stores or immature liver function. Newborns born preterm, with low birth weight, sepsis, hypothermia, hypoxic ischemic encephalopathy, polycythemia, or inadequate feeding are at particular risk. Newborns of diabetic mothers and those who are large for gestational age, with birth defects or with congenital metabolic diseases are also at risk. Symptoms of hypoglycemia, such as jitteriness, cyanosis, apnea, hypothermia, poor body tone, poor feeding, lethargy, and seizures, may be absent; therefore, blood glucose should be assessed routinely in these at-risk newborns. | (1) medication delivery mechanism (infusion kit + any IV fluid or single use syringes)<br>(2) gloves (disposable)<br>(3) soap and water for handwashing or alcohol-based hand rub<br>(4) disinfectant solutions (e.g. chlorine bleach)<br>(5) non-sharps waste container (pedal bin receptable with lid and plastic liner)<br>(6) sharps container<br>for heel prick:<br>(7) lancets<br>(8) alcohol swabs<br>(9) cotton wool | (1) 40% dextrose gel (oral)<br>(2) 10% glucose solution (IV) | (1) blood glucose – glucometer and glucometer test strips | (1) the health facility has written, up-to-date guidelines, protocols, and standard operating procedures for the care of small and sick newborns that include prevention and management of hypoglycemia in those at risk of impaired metabolic adaptation, consistent with who guidelines. | | | | | | |

| Intervention | Description of intervention | Equipment and supplies | Medicines and commodities | Diagnostics | Guidelines and staff training | Basic amenities | Routine service | Assessment | Intervention | Documentation and referral | Experience of care |
|---|---|---|---|---|---|---|---|---|---|---|---|
| | | | | | (2) staff trained in care of small and sick newborns that include prevention and management of hypoglycemia | | | | | | |
| Detection and management of jaundice (phototherapy) | Jaundice is yellow coloring of the skin and mucous membranes due to hyperbilirubinemia. Without treatment, it can lead to acute bilirubin encephalopathy (kernicterus), resulting in long-term motor, language, and hearing problems. Preterm and low-birth-weight newborns are at particular risk because of increased production of bilirubin and an immature liver. Most cases are treated with effective, safe phototherapy, preferably with high-intensity light emitting diodes (LED), and regular monitoring of blood bilirubin levels. | (1) eye patches/eye shields for baby (2) irradiance meter/spectro-radiometer (3) phototherapy lamps/units with fluorescent tubes (high intensity) or led phototherapy (4) spare fluorescent tubes (5) white linen for babies on phototherapy for cot and to cover unit (6) alcohol swabs (7) reusable probe tips (8) printed nomogram or (9) computer for patient monitoring (10) gloves (disposable) (11) soap and water for | (1) IV gamma globulin | (1) total serum bilirubin (TSB) (2) TCB (transcutaneous bilirubinometer or icterometer) (3) blood typing – abo blood grouping test (ABO grouping sera) (4) rhesus factor blood test (RH test sera, centrifuge) (5) Coombs test (Coombs reagent) (6) glucose-6- | (1) the health facility has written, up-to-date guidelines, protocols, and standard operating procedures for assessing and managing jaundice in newborns. (2) the health facility clinical staff who care for newborns receive training and regular refresher sessions in | (1) electricity supply | | | | | |

| Intervention | Description of intervention | Equipment and supplies | Medicines and commodities | Diagnostics | Guidelines and staff training | Basic amenities | Routine service | Assessment | Intervention | Documentation and referral | Experience of care |
|---|---|---|---|---|---|---|---|---|---|---|---|
| | | handwashing or alcohol based hand rub<br>(12) disinfectant solutions (e.g. chlorine bleach)<br>(13) non-sharps waste container (pedal bin receptable with lid and plastic liner)<br>(14) sharps container<br>for blood test:<br>(15) needle and syringe for blood draw | | phosphate dehydrogenase (g6pd)<br>(7) hemoglobin (full blood count, hematocrit or point-of-care hemoglobin) | assessing and managing newborns with jaundice | | | | | | |
| Detection and management of anemia | Anemia is defined as a hematocrit or hemoglobin concentration > 2 standard deviations below the mean for age. The main effects of anemia are reduced oxygen delivery to tissues, with clinical symptoms and acute or chronic consequences that include poor growth, decreased activity, limited cardiovascular reserve and adverse effects on cognitive development. Preterm newborns are at particular risk of anemia if they are born before placental iron transport and fetal erythropoiesis are complete, with low plasma levels of erythropoietin, both reduced production and accelerated catabolism and rapid neonatal growth. Red blood cell loss is increased in preterm infants by infection, bleeding, and | blood transfusion which requires:<br>(1) blood storage refrigerator functioning and temperature in required range for last 30 days<br>(2) neonatal blood transfusion set<br>(3) 4-way stopcock for umbilical venous line<br>(4) exchange transfusion sets<br>(5) portable monitors<br>(6) thermometer<br>(7) stethoscope<br>(8) gloves (disposable)<br>(9) soap and water for handwashing or alcohol based hand rub<br>(10) disinfectant solutions (e.g. | (1) iron (syrup)<br>(2) blood supply sufficiency (no interruption of blood availability in last three months)<br>(3) blood supply safety (blood obtained only from national or regional blood bank, or blood obtained from other sources but always screened for HIV, syphilis, hepatitis B, and hepatitis C) | (1) hemoglobin (full blood count, hematocrit, or point-of-care hemoglobin)<br>(2) blood typing – ABO blood grouping test (ABO grouping sera)<br>(3) cross-matching | (1) the health facility has written, up-to-date guidelines, protocols and standard operating procedures for assessing and managing anemia in small and sick newborns.<br>(2) the health facility clinical staff who care for small and sick | (1) electricity supply | | | | | |

| Intervention | Description of intervention | Equipment and supplies | Medicines and commodities | Diagnostics | Guidelines and staff training | Basic amenities | Routine service | Assessment | Intervention | Documentation and referral | Experience of care |
|---|---|---|---|---|---|---|---|---|---|---|---|
| | frequent blood sampling for laboratory testing. Small and sick newborns should be screened and managed for anemia and its complications and may require blood transfusion. | chlorine bleach)<br>(11) non-sharps waste container (pedal bin receptable with lid and plastic liner)<br>(12) sharps container<br><br>for blood test:<br>(13) needle and syringe for blood draw<br><br>for heel prick:<br>(14) lancets<br>(15) alcohol swabs<br>(16) cotton wool | | | newborns receive training and regular refresher sessions in assessing and managing anemia in small and sick newborns<br>(3) the health facility has written, up-to-date guidelines, protocols and standard operating procedures for safe blood transfusion for small and sick newborns, including indications for use, appropriate blood storage and monitoring before, during and after transfusion, and | | | | | | |

| Intervention | Description of intervention | Equipment and supplies | Medicines and commodities | Diagnostics | Guidelines and staff training | Basic amenities | Routine service | Assessment | Intervention | Documentation and referral | Experience of care |
|---|---|---|---|---|---|---|---|---|---|---|---|
| | | | | | transfusion reactions are managed and recorded. (4) staff trained in appropriate use of blood and safe blood transfusion | | | | | | |
| Detection and management of neonatal encephalopathy | Neonatal encephalopathy (NE) is defined as a condition occurring in babies born over 35 weeks gestational age in which there is disturbed neurological function. The key feature is the disturbance in the degree or quality of consciousness; other features, such as seizures, cardiorespiratory compromise or abnormal tone and reflexes, may occur alongside it but are not necessary to make the diagnosis. Knowledge gaps preclude a definitive test or set of markers that accurately identifies, with high sensitivity and specificity, an infant in whom NE is attributable to an acute intrapartum event. The term NE should be used where no definite etiological diagnosis is known, and hypoxic-ischemic encephalopathy (HIE) where clear diagnosis | (1) stethoscope (2) thermometer (3) hypoxic ischemic encephalopathy (HIE) scoring chart (4) gloves (disposable) (5) soap and water for handwashing or alcohol-based hand rub | | | (1) the health facility has written, up-to-date guidelines, protocols, and standard operating procedures for assessing and managing neonatal encephalopathy in newborns. (2) the health facility clinical staff who care for newborns receive training and regular | | | | | | |

| Intervention | Description of intervention | Equipment and supplies | Medicines and commodities | Diagnostics | Guidelines and staff training | Basic amenities | Routine service | Assessment | Intervention | Documentation and referral | Experience of care |
|---|---|---|---|---|---|---|---|---|---|---|---|
| | of hypoxia-ischemia is known to have led to the neonate's clinical state. | | | | refresher sessions in assessing and managing neonatal encephalopathy | | | | | | |
| Seizure management | Neonatal seizures are some of the most frequent neurological events in newborns, reflecting a variety of pre-, peri- or postnatal disorders of the central nervous system. They are also a common manifestation of metabolic abnormality in the newborn period and are often the first sign of neurological dysfunction. | (1) medication delivery mechanism (infusion kit + any IV fluid or single use syringes)<br>(2) gloves (disposable)<br>(3) soap and water for handwashing or alcohol-based hand rub<br>(4) non-sharps waste container (pedal bin receptable with lid and plastic liner)<br>(5) sharps container<br>(6) disinfectant solutions (e.g., chlorine bleach) | (1) phenobarbital (first line treatment) | | (1) the health facility has written, up-to-date guidelines, protocols, and standard operating procedures for assessing and managing seizures in newborns.<br>(2) the health facility clinical staff who care for small and sick newborns receive training and regular refresher sessions in assessing and managing | | | | | | |

| Intervention | Description of intervention | Equipment and supplies | Medicines and commodities | Diagnostics | Guidelines and staff training | Basic amenities | Routine service | Assessment | Intervention | Documentation and referral | Experience of care |
|---|---|---|---|---|---|---|---|---|---|---|---|
| | | | | | seizures in newborns | | | | | | |
| Safe administration of intravenous fluids | All small and sick newborns should be provided with supportive care and closely monitored. Small and sick newborns who require supportive care, such as intravenous fluids, should be given these only when indicated. Attention to safe use of intravenous fluids is particularly important for small and sick newborns. Medication doses and fluid volumes are based on the newborn's weight, and an incorrect weight or measurement error can lead to administration of an inappropriate dose or volume. Medications are administered as per standard procedures to avoid serious adverse events. The volumes and rates of administration of intravenous fluids must be strictly monitored to avoid fluid overload. | (1) infant scale<br>(2) butterfly sets (22-25 gauge) or<br>(3) sterile needles (19-26 gauge)<br>(4) IV tubing/infusion set (neonatal giving set) with burette 100-150ml, sterile, single use<br>(5) IV infusion stands on castors<br>(6) sterile syringes (small sizes 0.5, 1ml, 2ml, 5ml, 10ml, 20ml)<br>(7) stopcocks 2 or 3 way<br>(8) syringe driver/syringe pumps 10, 20, 50ml (single phase)<br>(9) non-sharps waste container (pedal bin receptable with lid and plastic liner)<br>(10) disinfectant solutions (e.g. chlorine bleach)<br>(11) gloves (disposable)<br>(12) sharps container<br>(13) soap and water for handwashing or alcohol-based hand rub | (1) sodium chloride 0.9%<br>(2) ringer's lactate<br>(3) 5% dextrose – normal saline | | (1) the health facility has written, up-to-date guidelines, protocols and standard operating procedures for safe use of intravenous fluids and parenteral nutrition in selected critically ill neonates.<br>(2) the health facility has instructions for newborn-specific dosage and charts and electronic calculators to assist health professionals who care for small and sick newborns in | (1) separate area/clean space for preparing IV fluids (can be the same area as for preparation of IV drugs) | | | | | |

| Intervention | Description of intervention | Equipment and supplies | Medicines and commodities | Diagnostics | Guidelines and staff training | Basic amenities | Routine service | Assessment | Intervention | Documentation and referral | Experience of care |
|---|---|---|---|---|---|---|---|---|---|---|---|
| | | | | | selecting and administering correct doses of intravenous fluids and medication to newborns. (3) the health facility has written, up-to-date guidelines, protocols and standard operating procedures for the prevention, monitoring, detection and management of extravasation tissue injuries resulting from infusions. (4) health professionals receive in-service training and regular refresher | | | | | | |

| Intervention | Description of intervention | Equipment and supplies | Medicines and commodities | Diagnostics | Guidelines and staff training | Basic amenities | Routine service | Assessment | Intervention | Documentation and referral | Experience of care |
|---|---|---|---|---|---|---|---|---|---|---|---|
| | | | | | sessions on newborn monitoring and supportive care | | | | | | |
| Detection and referral management of birth defects | There are many types of congenital anomaly, but only a few are common. Some require urgent surgical attention, while others should be left until the child is older. Early recognition results in better outcomes and allows the parents to inform themselves about treatment options. The majority of congenital abnormalities are detectable at birth, and two thirds of deaths due to these conditions are preventable with pediatric surgery and neonatal intensive care. The most common congenital abnormalities are cleft lip and palate, congenital heart anomalies and neural tube defects (spina bifida). | | | | (1) the health facility has written, up-to-date guidelines, protocols, and standard operating procedures for assessing and managing newborns with congenital abnormalities, including for specialist consultation and referral pathways. | (1) emergency transportation | | | | | |

### *Table S1: Detailed definitions for maternal PNC index items*

| Sub-domain | Item | Detailed indicator definition | Data availability notes | Suggested calculation notes |
|---|---|---|---|---|
| Equipment and supplies | Blood pressure apparatus | Either manual or digital blood pressure apparatus observed available and functional in the delivery or OPD service provision area. | Only collected in OPD for SARA | |
| Equipment and supplies | Stethoscope | Stethoscope observed available and functional in the delivery or OPD service provision area. | Only collected in OPD for SARA | |
| Equipment and supplies | Thermometer | Thermometer observed available and functional in the delivery or OPD service provision area. | Only collected in OPD for SARA | |
| Equipment and supplies | Sputum collection container | Facilities that offer TB services have sputum collection container observed available in the TB service area. | Not collected in SARA | |
| Equipment and supplies | Single-use standard disposable syringe with needles or auto-disable syringes with needle | Facilities that offer HIV diagnostic services have single-use standard disposable syringe with needles or auto-disable syringes with needle observed available in the delivery, OPD, HIV service provision area or in general storage. | Not collected in SARA | |
| Equipment and supplies | Environmental disinfectant | Disinfectant (e.g., chlorine, hibitane, alcohol) is observed available in the delivery service area, OPD service provision area, HIV service area if the facility offers HIV testing services, TB service area if the facility offers TB testing services, or general storage. | Only collected in OPD for SARA | |
| Equipment and supplies | Clean/sterile gloves | Disposable latex gloves are observed available in the delivery service area, OPD service provision area, HIV service area if the facility offers HIV testing services, TB service area if the facility offers TB testing services, or general storage. | Only collected in OPD for SARA | |
| Equipment and supplies | Non-sharps waste | Non-sharps waste (pedal bin receptable with lid and plastic liner) is observed in delivery service area, OPD service provision area, HIV service area if the facility offers HIV testing services, and TB service area if the facility offers TB testing services. | Only collected in OPD for SARA | |
| Equipment and supplies | Sharps container | Facilities that offer HIV diagnostic services have a sharps container that is observed in delivery service area, OPD service provision area, or HIV service area. | Only collected in OPD for SARA | |
| Equipment and supplies | Handwashing materials | Facility has either handwashing soap and running water or hand sanitizer observed available in delivery service area, OPD service provision area, HIV service area if the facility offers HIV testing services, and TB service area if the facility offers TB testing services. | Only collected in OPD for SARA | |
| Equipment and supplies | Examination light | Examination light or flashlight, is observed available and functioning in the delivery or OPD service provision area. | | |

| Sub-domain | Item | Detailed indicator definition | Data availability notes | Suggested calculation notes |
|---|---|---|---|---|
| Guidelines | Guidelines containing information on pregnancy (e.g., IMPAC) | Guidelines containing information on pregnancy (e.g., IMPAC) are observed available within the delivery service provision area. | | |
| Medicines and commodities | Supplement containing iron | Iron containing supplements observed at least one non-expired in ANC service provision area or general storage. | SPA collects data on stand-alone iron tablets as well as iron+ folic acid combination tablets. SARA only collects data on stand-alone iron tablets. | |
| Medicines and commodities | Albendazole or mebendazole | Albendazole or mebendazole observed at least one non-expired in general storage. | | |
| Medicines and commodities | PrEP | Facilities that offer HIV diagnostic services have at least one non-expired Tenofovir disoproxil fumarate (TDF) or Tenofovir + Emtricitabine [TDF + FTC] in general storage. | Collected only for countries with substantial HIV burden | |
| Medicines and commodities | Mix of FP methods | A mix of FP methods (i.e., one short-acting (combined injectable contraceptives, combine oral contraceptive pills, emergency contraceptive pills, progesterone only contraceptive pills, progesterone only injectable contraceptive), one long-acting or permanent (implants, or IUD), one barrier or non-hormonal methods (female condom or male condoms) is available in general storage. | | |
| Medicines and commodities | Paracetamol | Paracetamol observed at least one non-expired in general storage. | | |
| Diagnostics | HIV diagnostic capacity | Facility reported that it had the capacity to conduct HIV testing in the facility, either by rapid diagnostic testing or ELISA, and an unexpired HIV rapid diagnostic test kit was observed to be available in the facility on the day of the survey, or dynabeads test with vortex mixer was observed to be available in the facility on the day of the visit, or western blot test was observed to be available in the facility on the day of the visit. | | |
| Diagnostics | TB diagnostic capacity | Facility reported that it had the capacity to conduct TB testing in the facility by microscopy, where either light or fluorescent microscope, slides, and ZN stain; or fluorescent microscope, slides and auramine-rhodamine stain was observed to be available in the facility on the day of the survey, or facility has an observed system for sending sputum outside facility for TB diagnosis and receiving results. | | |

| Sub-domain | Item | Detailed indicator definition | Data availability notes | Suggested calculation notes |
|---|---|---|---|---|
| Basic amenities | Room is private room with auditory and visual privacy | Private room with auditory and visual privacy is observed in delivery, OPD service provision area, or HIV service area if the facility offers HIV testing services. | Only collected in OPD for SARA | |
| Staff training | Training in IMPAC | Proportion of health care workers delivering newborn care services that have been trained in the last two years in pregnancy (i.e., IMPAC). | | |
| Staff training | Training in early and exclusive breastfeeding | Proportion of health care workers delivering newborn care services that have been trained in the last two years in early and exclusive breastfeeding. | | |
| Staff training | Available HIV guidelines and staff training | Proportion of health care workers delivering newborn care services that have been trained in the last two years in providing counseling related to HIV testing and conducting HIV tests; and facilities have guidelines containing information on HIV counselling and testing observed available within the HCT service provision area; and national ART guidelines are observed available within the HIV service provision area. | | 3 variables were created: (1) proportion of staff trained in counseling related to HIV testing and conducting HIV test; (2) availability of HIV testing guidelines; and (3) availability of ART guidelines<br><br>The mean of the 3 variables was calculated |
| Staff training | Available TB guidelines and staff training | Proportion of health care workers delivering newborn care services that have been trained in the last two years in diagnosis of tuberculosis based on sputum tests or analysis; and facilities have national guidelines on TB diagnosis and treatment observed available within the TB service provision area. | | 2 variables were created: (1) proportion of staff trained in diagnosis of tuberculosis based on sputum tests or analysis; and (2) availability of national TB guidelines<br><br>The mean of the 2 variables was calculated |

*Table S2: Detailed definitions for newborn PNC index items*

| Sub-domain | Item | Detailed indicator definition | Data availability notes | Suggested calculation notes |
|---|---|---|---|---|
| Equipment and supplies | Stethoscope | Stethoscope observed available and functional in the delivery or OPD service provision area. | Only collected in OPD for SARA | |
| Equipment and supplies | Thermometer | Thermometer observed available and functional in the delivery or OPD service provision area. | Only collected in OPD for SARA | |
| Equipment and supplies | Single-use standard disposable syringe with needles or auto-disable syringes with needle | Single-use standard disposable syringe with needles or auto-disable syringes with needle observed available in the delivery, child vaccination or OPD service provision area or in general storage. | Only collected in vaccination for SARA | |
| Equipment and supplies | Vaccine documentation | Blank/unused individual child vaccination cards or booklets, and immunization tally sheet observed in child vaccination service provision area. | Not collected in SARA | Scored as 0/0.5/1 |
| Equipment and supplies | Refrigerator with temperature monitoring device and power, or vaccine carrier with ice packs | Refrigerator with temperature monitoring device and power (central supply, generator with fuel or battery, or solar) is observed in facility; or vaccine carrier with ice packs is observed in facility. | Temperature monitoring device not collected in SARA | |
| Equipment and supplies | Environmental disinfectant | Disinfectant (e.g., chlorine, hibitane, alcohol) is observed available in the delivery, child vaccination or OPD service provision area, or general storage. | Only collected in OPD for SARA | |
| Equipment and supplies | Clean/sterile gloves | Disposable latex gloves observed available in the delivery, child vaccination, or OPD service provision area, or general storage. | Only collected in OPD for SARA | |
| Equipment and supplies | Non-sharps waste | Non-sharps waste (pedal bin receptable with lid and plastic liner) is observed in delivery, child vaccination, or OPD service provision area. | Only collected in OPD for SARA | |
| Equipment and supplies | Sharps container | Sharps container observed available in the delivery, child vaccination or OPD service provision area. | Only collected in OPD for SARA | |
| Equipment and supplies | Handwashing materials | Facility has either handwashing soap and running water or hand sanitizer observed available in the delivery, child vaccination or OPD service provision area. | Only collected in OPD for SARA | |
| Basic amenities | Room is private room with auditory and visual privacy | Private room with auditory and visual privacy is observed in the delivery or OPD service provision area. | Only collected in OPD for SARA | |
| Guidelines | Guidelines containing information on newborn care (e.g., IMPAC or ENC) | Guidelines containing information on newborn care (e.g., IMPAC or ENC) are observed available within the delivery service provision area. | | |

| Sub-domain | Item | Detailed indicator definition | Data availability notes | Suggested calculation notes |
|---|---|---|---|---|
| Guidelines | Child vaccination guidelines | Guidelines containing information on child vaccination are observed available within the delivery service provision area. | | |
| Medicines and commodities | Chlorhexidine solution | In countries with a chlorhexidine policy, chlorhexidine solution observed in pharmacy or anywhere in the facility where medicines are routinely stored; at least one with valid expiration date. | Not collected in SARA | |
| Medicines and commodities | Child vaccinations | BCG, HEPB, and OPV0 observed in pharmacy or anywhere in the facility where medicines are routinely stored; at least one with valid expiration date. | | Calculated as 0.33, 0.66, 1 depending on availability of each of the vaccines |
| Staff training | Training in IMPAC or newborn care | Proportion of health care workers offering delivery or newborn care services that have been trained in newborn care (i.e., IMPAC; thermal care; early and exclusive breastfeeding; and sterile cord cutting and appropriate cord care) in the last two years. | SARA only collects IMPAC training and not on specific ENC topics | Each individual was coded as either having received IMPAC training (1 point) or newborn care training (0, 0.33, 0.66, 1 points based on having training in 3 topics- early and exclusive breastfeeding, thermal care and sterile cord cutting and appropriate care). Each individual received whichever score was higher. Scores were then converted to the proportion of staff trained. |

### *Table S3: Detailed definitions for small and sick newborn care index items*

| Sub-domain | Item | Detailed indicator definition | Data availability notes | Suggested calculation notes |
|---|---|---|---|---|
| **Immediate newborn care and Routine care** | | | | |
| Equipment and supplies | Linen for drying baby | Linen for wrapping and drying the newborn observed available in the delivery service provision area. | Not collected in SARA; collected for some SPAs | |
| Equipment and supplies | Cord cutting supplies | Sterile scissors/blade to cut cord and cord clamp OR sterile delivery pack observed available in the delivery service provision area. | | |
| Equipment and supplies | Thermometer for low-body temperature | Thermometer for low-body temperature observed available and functional in the delivery service provision area. | Not collected in SARA; collected for some SPAs | |
| Medicines and commodities | Vitamin K | Vitamin K observed in pharmacy or anywhere in the facility where medicines are routinely stored; at least one with valid expiration date observed available. | Not collected in SARA; collected for some SPAs | |
| Medicines and commodities | Antibiotic eye ointment (tetracycline or other) | Antibiotic eye ointment for newborns observed in pharmacy or anywhere in the facility where medicines are routinely stored; at least one with valid expiration date observed available. | | |
| Medicines and commodities | Chlorhexidine solution | Chlorhexidine solution observed in pharmacy or anywhere in the facility where medicines are routinely stored; at least one with valid expiration date observed available. Only applies to countries where chlorhexidine use is included in national policy. | Not collected in SARA | |
| Staff training | Staff trained in clean cord cutting and appropriate cord care | Proportion of health care workers delivering newborn care services that have been trained in clean cord cutting and appropriate cord care in the last two years. | Not collected in SARA | |
| Staff training | Staff trained in thermal care | Proportion of health care workers delivering newborn care services that have been trained in thermal care (including immediate drying and skin-to-skin care) in the last two years. | Not collected in SARA | |
| Equipment and supplies | Radiant heater/warmth source | External heat source (other than incubator) observed available and functional in the delivery service provision area. | | |
| Routine services | Routine, complete (head-to-toe) examination of newborn before discharge | Facility practices routine, complete (head-to-toe) examination of newborn before discharge | Not collected in SARA | |
| Medicines and commodities | Immunization supplies | 1) BCG, 2) Hep B, and 3) OPV observed in pharmacy or anywhere in the facility where medicines are routinely stored; at least one with valid expiration date observed available.<br>4) Single use needles and syringes are observed available in the delivery service provision area, OPD, pharmacy, or immunization service area. | | Calculated as a percentage of items available (total of 6 items possible); score ranges from 0 to 1 |

| Sub-domain | Item | Detailed indicator definition | Data availability notes | Suggested calculation notes |
|---|---|---|---|---|
| | | 5) Guidelines containing information on child immunization or the national immunization schedule are observed available in the immunization service provision area.<br>6) A refrigerator dedicated for vaccine storage with a temperature monitoring device and consistent electricity supply is available in the facility OR the facility has a cold box/vaccine carrier with ice packs in the immunization service area. | | |
| **Early initiation and support for breastfeeding** | | | | |
| Staff training | Staff trained in early and exclusive breastfeeding | Proportion of health care workers delivering newborn care services that have been trained in early and exclusive breastfeeding in the last two years. | Not collected in SARA | |
| Routine services | Initiation of breastfeeding within the first hour | Facility routinely practices initiation of breastfeeding within the first hour. | Not collected in SARA | |
| **Neonatal resuscitation** | | | | |
| Equipment and supplies | Airway suction apparatus (suction apparatus with catheter or suction bulb for mucus extraction) | Airway suction apparatus (suction apparatus with catheter or suction bulb for mucus extraction) observed available and functional in the delivery service provision area. | | |
| Equipment and supplies | Infant resuscitation bag/mask | Infant resuscitation bag/mask observed available and functional in the delivery service provision area. | | |
| Staff training | Staff trained in neonatal resuscitation using bag and mask | Proportion of health care workers delivering newborn care services that have been trained in neonatal resuscitation using bag and mask in the last two years.<br>Alternative: At least one health care worker providing delivery services has been trained in neonatal resuscitation using bag and mask in the last two years. | | |
| Routine services | Facility past three months provided neonatal resuscitation | Providers have carried out neonatal resuscitation as part of their work in the facility in the past three months. | Not collected in SARA | |
| **Prevention of mother-to-child transmission of HIV (PMTCT)** | | | | |
| Basic amenities | PMTCT room is private room with auditory and visual privacy | PMTCT room is private room with auditory and visual privacy in the delivery or PMTCT service provision areas. | | |
| Medicines and commodities | Antiretrovirals for newborns | Antiretrovirals for babies (NVP syrup or AZT syrup only) observed in pharmacy or anywhere in the facility where medicines are routinely stored; at least one with valid expiration date. | | |
| Medicines and commodities | Antiretrovirals for mothers | Antiretrovirals for mothers (country-specific first line ARV prophylaxis for HIV-positive pregnant women) observed in pharmacy or anywhere in the facility where | | |

| Sub-domain | Item | Detailed indicator definition | Data availability notes | Suggested calculation notes |
|---|---|---|---|---|
| | | medicines are routinely stored; at least one with valid expiration date. | | |
| Medicines and commodities | Cotrimoxazole (preventative therapy) | Cotrimoxazole syrup observed in pharmacy or anywhere in the facility where medicines are routinely stored; at least one with valid expiration date. | | |
| Diagnostics | HIV diagnostic capacity | Facility has the ability to conduct quantitative nucleic acid testing for HIV diagnosis (PCR) or RNA / PCR for DNA-EID + filter paper for DBS OR needle and syringe for blood draw; needle and syringe can be in PMTCT, OPD, delivery, or general supplies areas | | |
| Guidelines | PMTCT guidelines and Infant and young child feeding counseling guidelines | Guidelines for PMTCT are available in the delivery or PMTCT service provision areas. | | Calculated as a percentage of items available (total of 2 items possible); score ranges from 0 to 1 |
| | | Guidelines for infant and young child feeding are observed available in the delivery or PMTCT service provision areas. | | |
| Staff training | Staff trained in ARV prophylactic treatment for PMTCT | Proportion of health care workers delivering PMTCT services that have been trained in PMTCT or ARV prophylactic treatment for PMTCT in the last two years.<br>Alternative: At least one health care worker providing PMTCT services has been trained in PMTCT in the last two years. | | |
| Staff training | Staff trained in newborn nutrition counseling of mother with HIV or infant and young child feeding | Proportion of health care workers delivering PMTCT services that have been trained in newborn nutrition counseling of mother with HIV OR infant and young child feeding in the last two years.<br>Alternative: At least one health care worker providing PMTCT services has been trained in newborn nutrition counseling of mother with HIV OR infant and young child feeding in the last two years. | | |
| **Kangaroo mother care (KMC)** | | | | |
| Basic amenities | Separate room or space for KMC | Facility has a separate room or space for KMC. | Not collected in SARA | |
| Staff training | Staff trained in KMC for low-birth-weight babies | Proportion of health care workers delivering newborn care services that have been trained in KMC for low-birth-weight babies in the last two years. | Not collected in SARA | |
| Routine services | Facility practices KMC | Facility practices KMC. | Not collected in SARA | |
| **Detection and management of neonatal infection** | | | | |
| Medicines and commodities | Antibiotic treatment for neonatal infection | Antibiotic treatment for neonatal infection (Outpatient treatment: Gentamycin (IM or IV) and Amoxicillin (syrup/oral suspension) OR Inpatient treatment: (Benzylpenicillin (IV or IM) OR Ampicillin (IV or IM)) AND Gentamycin (IM or IV)) observed in pharmacy or anywhere in the facility where medicines are routinely stored; at least one with valid expiration date. | | |

| Sub-domain | Item | Detailed indicator definition | Data availability notes | Suggested calculation notes |
|---|---|---|---|---|
| Staff training | Staff trained in newborn infection management | Proportion of health care workers delivering newborn care services that have been trained in newborn infection management (including injectable antibiotics) in the last two years. | Not collected in SARA | |
| Diagnostics | Full blood count | Facility has the ability to conduct hemoglobin testing + functioning hematology analyzer or functioning blood chemistry analyzer available and functioning | | |
| Diagnostics | Chest x-ray | Facility has the ability to conduct diagnostic x-ray testing + digital x-ray machine (not requiring film) or x-ray machine and unexpired film available and functioning | | |
| **Comfort and pain management** | | | | |
| Medicines and commodities | Paracetamol | Paracetamol syrup observed in pharmacy or anywhere in the facility where medicines are routinely stored; at least one with valid expiration date. | | |
| Medicines and commodities | Morphine | Morphine (syrup or injectable) observed in pharmacy or anywhere in the facility where medicines are routinely stored; at least one with valid expiration date. | Not collected in SPA; country specific addition | |
| **Detection and management of hypoglycemia** | | | | |
| Diagnostics | Blood glucose testing | Facility has the ability to conduct blood glucose testing + glucometer and test strips available and functioning or blood chemistry analyzer available and functioning | | |
| Medicines and commodities | Glucose injectable solution | Glucose injectable solution observed in pharmacy or anywhere in the facility where medicines are routinely stored; at least one with valid expiration date. | | |
| **General readiness items** | | | | |
| Equipment and supplies | Thermometer | Thermometer observed available and functional in the delivery service provision area or OPD. | | |
| Equipment and supplies | Infant scale | Infant weighing scale observed available and functional in the delivery service provision area or OPD. | | |
| Equipment and supplies | Stethoscope | Stethoscope observed available and functional in the delivery service provision area. | | |
| Equipment and supplies | Medication delivery mechanism (infusion kit + any IV fluid OR single use syringes) | Medication delivery mechanism (pediatric infusion kit + any IV fluid observed anywhere in the facility where supplies are routinely stored OR single use syringes observed in the delivery service provision area). For IV fluids, observed at least one with valid expiration date. | | |
| Equipment and supplies | Pulse oximeter | Pulse oximeter observed available and functional in the OPD. | | |
| Equipment and supplies | Oxygen supply | Oxygen cylinders, concentrators, or an oxygen distribution system observed available and functional in the OPD, and consistent electricity supply is available in the facility. | | |

| Sub-domain | Item | Detailed indicator definition | Data availability notes | Suggested calculation notes |
|---|---|---|---|---|
| Equipment and supplies | Non-sharps waste container | Pedal bin waste receptacle with lid and plastic liner observed available in the delivery service provision area. | | |
| Equipment and supplies | Environmental disinfectant | Disinfectant (e.g., chlorine, hibitane, alcohol) is observed available in the delivery service provision area or OPD. | | |
| Equipment and supplies | Clean/sterile gloves | Disposable latex gloves are observed available in the delivery service provision area. | | |
| Equipment and supplies | Sharps container | Sharps container observed available in the delivery service provision area. | | |
| Equipment and supplies | Handwashing supplies | Soap and running water OR alcohol-based hand rub observed available in the delivery service provision area. | | |
| Guidelines | Guidelines containing information on newborn care (e.g., ENC, IMPAC, BEmOC, CEmOC) | Guidelines containing information on newborn care (e.g., ENC, IMPAC, BEmOC, CEmOC) are observed available in the delivery service provision area. | | |
